# Reducing sedentary behaviour in patients after stroke (RECREATE): exploratory economic evaluation from an external pilot cluster randomised trial of the ‘Get Set Go’ intervention

**DOI:** 10.1101/2025.10.18.25338286

**Authors:** Anita Patel, Nahel Yaziji, Barbara Barrett, Renee Romeo, Bethan Copsey, Lauren Moreau, Florence Day, Alexandra Welsh, Jennifer Airlie, Seline Ozer, Louisa-Jane Burton, Jessica Johansson, Claire Fitzsimons, Gillian Mead, Amanda Farrin, Anne Forster, the RECREATE PMG

## Abstract

**Background:** Sedentary behaviour is common among stroke survivors and contributes to poor long-term health outcomes and high societal costs. The *Get Set Go (GSG)* intervention was developed to reduce sedentary behaviour through a whole-service approach spanning inpatient and community stroke care. This paper reports an exploratory economic evaluation conducted alongside the external pilot cluster randomised trial of GSG within the RECREATE programme.

**Objectives:** The evaluation examined the potential cost-effectiveness of GSG compared to usual care and assessed the feasibility of alternative methods for capturing economic data. Specific aims included estimating costs and outcomes from NHS/personal social services (PSS) and societal perspectives, assessing the agreement between self-reported and routinely collected Hospital Episode Statistics (HES) data, and exploring the implications of different data sources and follow-up periods for future evaluations.

**Methods:** Stroke services across England and Scotland were randomised (eight intervention, seven control), recruiting 334 stroke survivors. Costs were derived from self-reported service use (Client Service Receipt Inventory) and HES data, and outcomes were measured using the Nottingham Extended Activities of Daily Living Scale (NEADL) and EQ-5D-5L-derived quality-adjusted life years (QALYs) over 12 and 24 months. Analyses followed an intention-to-treat approach using bootstrap mixed-effects models.

**Results:** From both NHS/PSS and societal perspectives, mean costs were higher and outcomes (QALYs and NEADL) were lower in the GSG arm compared to usual care, suggesting that GSG was unlikely to be cost-effective. Self-reported data underestimated secondary care use compared with HES, producing lower total cost estimates. While HES-derived costs were higher overall, they did not replicate the higher costs observed for GSG using self-report data. Substantial missing data and reduced follow-up due to the COVID-19 pandemic limited interpretation of long-term cost-effectiveness.

**Conclusions:** This exploratory analysis indicates that GSG is unlikely to be cost-effective and highlights methodological lessons for future trials. Routine data, such as HES, may provide a more reliable and less burdensome source for estimating secondary care costs, whereas self-report remains important for capturing unpaid care and community-based resource use. A hybrid data collection approach is recommended for future economic evaluations in stroke rehabilitation.

## INTRODUCTION

There is increasing evidence that sedentary behaviour has a detrimental effect on health and wellbeing. At least 1.2 million people living in England have had a stroke, and longer-term outcomes are poor for many. Associated societal costs for the United Kingdom (UK) are estimated at £26 billion annually [1] and are set to treble by 2035 [2]. Stroke survivors are particularly sedentary compared to age-matched controls and addressing such behaviour may help improve longer term outcomes and thus alleviate some of these substantial costs.

The RECREATE research programme sought to enhance health outcomes for stroke survivors through the development and evaluation of strategies to reduce sedentary behaviour. Research questions for the programme related to: identifying existing evidence; exploring capabilities, opportunities and motivation related to sedentary behaviour after stroke; determining whether an intervention developed using principles of co-production was feasible to implement and evaluate; and assessing the clinical and cost-effectiveness of the intervention – we report here the evaluation related to this last question.

A pragmatic definitive trial to assess the effectiveness and cost-effectiveness of the developed intervention (‘Get Set Go’ (GSG)) was planned and initiated. However, the overall aims changed part way through due to delays in site and participant recruitment caused by the COVID-19 pandemic. Consequently, the trial was reduced to an external pilot trial and the economic evaluation goals moved away from assessing the cost-effectiveness of GSG, to instead undertaking an economic evaluation on an exploratory basis only and addressing additional feasibility-related questions to support future research in similar patient groups. This paper reports the results of the exploratory economic evaluation of GSG. Results of the effectiveness and process evaluation are reported elsewhere [3].

## METHODS

### Overall trial design

The study was originally planned as a definitive multi-centre cluster-randomised trial with stroke survivors, with embedded process and economic evaluations. Conversations with interested stroke services began in October 2019. While the previous workstreams of the research programme had progressed according to planned timelines, the start of the definitive trial was disrupted by the COVID-19 pandemic. Following discussions with the Programme Steering Committee and funder (NIHR), we amended the design to a large external pilot trial to keep to original timelines. The amended trial aimed to recruit 300-400 participants across 15 stroke services. Recruitment of stroke survivors and carers started in January 2021. Further details of participant eligibility and exclusion etc. is available in the protocol for the external pilot trial [4].

Fifteen NHS stroke services in the UK (acute and/or rehabilitation stroke unit with a linked community service over a defined geographical area) were randomised on a 1:1 basis to deliver GSG alongside usual care (n=8) or usual care alone (n=7). Usual care was as determined by local policy and practices. Randomisation was stratified by Sentinel Stroke National Audit Programme (SSNAP) [5] grade, stroke unit composition and stroke unit size. The primary objective was to undertake a preliminary exploration of whether Get Set Go is likely to improve the ability to complete extended activities of daily living (EADLs) in the first year after stroke. A key secondary objective was to explore whether the intervention reduces sedentary behaviour. Assessments and outcome measures were undertaken at baseline and 6– and 12-months following registration; a subset of participants also completed outcome measures at 24-months post-registration.

### Intervention and training

An intervention to reduce sedentary behaviour was co-produced in the earlier stages of the research programme, informed by systematic reviews of quantitative and qualitative evidence related to sedentary behaviour, and observations and semi-structured interviews in two stroke services which explored the capability, opportunity and motivation to address sedentary behaviours. A case study and action research approach in three services was used to test implementation and clarify trial procedures and contributed to iterative refinement of the intervention. The resulting intervention (called Get Set Go (GSG)) was a whole-service intervention initiated in the inpatient stroke unit setting and continuing post-discharge for at least 12 weeks. The intervention focused on: (1) educating staff and stroke; (2) preparing and enabling staff to support and encourage stroke survivors to stand and move more in everyday stroke care; (3) encouraging stroke survivors to monitor their own standing and moving,

Intervention training with staff took place over a 24-month period (February 2021-February 2023) and comprised 103 training sessions delivered to 381 staff, primarily remotely (86.4%) due to COVID-19 restrictions, supported by in-person visits when allowed. Median session length was 60 minutes, and the median number of attendees was 4 (with a maximum of 23 attendees) per session.

### Economic evaluation objectives

The revised economic evaluation sought to address the following questions:

1. How likely is the GSG intervention to be cost-effective?
2. What is the difference in estimates for the cost of hospital services utilisation using data from self-report versus routinely collected hospital episode statistics (HES)?
3. What is the impact on cost-effectiveness estimates if the economic evaluation is based on self-report versus HES data?
4. What is the agreement level between estimates of hospital services utilisation based on HES versus self-reported data?
5. What additional information does a longer term (24m versus 12m versus 6m) follow up provide for the evaluation in terms of cost difference and completion rates?
6. Can HES data be used as a replacement for self-report service use data for study participants for whom data are missing?

The originally planned within-trial economic evaluation therefore proceeded but on an exploratory, rather than definitive basis, and served as a base case against which to consider the additional feasibility-related questions. Undertaking the economic evaluation thus remained a significant portion of the planned work and methods for this are detailed below. In brief, we sought to undertake exploratory cost-effectiveness and cost-utility analyses based on measures of resource use and outcomes over a 12– and 24-month follow-up period from the point of randomisation.

### Perspective, and identification and measurement of resource use data

The primary evaluation perspective was NHS and personal social services (NHS/PSS). This included costs associated with the intervention, secondary care, and primary/community-based health and social care. An additional societal perspective further added costs associated with non-NHS community-based services (where costs were borne by the patient) and unpaid help to the total NHS/PSS costs. Relevant resources to measure (see Supplementary Appendix, Box S1) were identified based on two previous large stroke rehabilitation trials [6, 7] and discussions with members of the study team and the Trial Steering Committee (TSC) which included patient representatives. We measured non-intervention resources using an adapted version of a Client Service Receipt Inventory (CSRI) which we developed and used in those previous trials. This was administered as a participant self-reported questionnaire, as part of the trial case report form, at baseline (covering the use of resources over the previous three months), at 6-months (for the period since baseline), at 12-month and 24-month follow ups (covering the previous 6 months); respondents were asked to report the frequency of use for each of the afore-listed resources within these assessment periods.

Use of inpatient, day patient, and outpatient services derived from Hospital Episode Statistics (HES) were available for the whole follow-up period, and are split between 01-12 months, and 12-24 months.

### Intervention costs, including training activities

Considering the complex nature of GSG, a mixed methods strategy was used to estimate its costs. Some cost elements were general across the intended patient population and were therefore estimated at a higher system level and then allocated to individual participants in equal proportion (i.e. ‘top-down approach’), while other elements were calculated based on specific inputs provided to each participant (i.e. ‘bottom-up approach’). In essence, fixed or semi-fixed costs were assigned to staff training time and management activities, and variable costs for activities related to staff interactions with participants.

Management comprised time spent in initial meetings with local PIs and the implementation groups where plans for delivering the training and the intervention were discussed. Training costs were estimated using data about initial and refresher training that were completed in the period prior to the service opening to recruitment, in addition to ongoing training during the implementation and delivery phase. We developed a detailed proforma for each intervention site to use to record the time spent providing training and the time spent delivering the intervention. We generated two sets of data collection for delivery costs, one for the inpatient setting and another for the community setting. Intervention delivery costs were then estimated at the participant level using data collected at the initial assessment and re-assessment of the participant’s traffic light status and the time spent prescribing the GSG approach. We also collated information about the number of personnel involved and their average pay grades. Resulting unit cost estimates for the intervention are summarised in Table 1.

**Table 1:** Estimated resources and costs of the intervention.

| Table 21. Estimated resources and costs of the intervention |  |  |
| --- | --- | --- |
| Delivering/receiving training |  |  |
| Mean (SD) time in minutes | 53 (51) | Time spent delivering initial and refresher training that was completed in the period prior to the service opening to recruitment, in addition to ongoing training during the implementation and delivery phase. |
| Overall number of staff who participated across all study sites | 822 |  |
| Number of training sessions delivered across all study sites | 179 |  |
| Mean (SD) number of staff per training session | 5 (3.4) |  |
| Delivering the intervention |  |  |
| Mean (SD) time in minutes assessing patient using the traffic light system | 11 (10.7) | Time spent delivering the intervention at the participant level using data collected at the initial assessment and re-assessment of the patient's traffic light status and the time spent prescribing the Get Set Go approach. |
| Mean (SD) time in minutes spent delivering the GSG, as per GSG therapy records | 27 (15.5) |  |
| Mean (SD) time in minutes spent delivering the GSG, as per inpatient GSG therapy records | 8 (8.1) |  |
| Mean (SD) time in minutes spent delivering the GSG, as per community GSG therapy records | 5 (3.9) |  |
| Costs |  |  |
| Total (average per participant) cost of management and planning activities<sup>1</sup> | £1457 (£8) | Based on quantity of time spent in planning, the number of personnel involved, and the average pay grade (pay grade ranged from band 6 (£53 per hour) to band 8 (£72 per hour)). |
| Total (average per participant) cost of training<sup>2</sup> | £44704 (£247) | Using information about length of training, professional qualifications of those involved in delivering and receiving the training, grade (pay grade ranged from band 2 (£30 per hour) to band 8 (£72 per hour), and the number of whole-time equivalent staff. |
| Total (average per participant) cost of delivering the intervention | £5323 (£29) | Using information about time spent in assessment/prescribing, staff profession, and most recent AfC band/pay band including spine point (pay grade ranged from band 2 (£30 per hour) to band 8 (£72 per hour)). |
| Total (average per participant) intervention cost | £51483 (£284) |  |

1. Based on 30 professional personnel, median pay-grade is 7, average meeting time per site is 45 minutes

### Costing resource utilisation

Relevant unit costs were applied to estimate individual-level costs for each timepoint. Unit cost values were derived from the relevant annual Personal Social Services Research Unit (PSSRU) unit cost compendium, National Health Service Reference Costs or other published sources (see Table 2 for details). Unpaid help was costed using an opportunity cost approach based on national average earnings and assuming 36.5 working hours per week. To estimate the costs of NHS professionals’ time (including staffing inputs for GSG) the relevant Agenda for Change pay bands were applied, with additional adjustments for staff overheads and salary “on-costs” as outlined in the PSSRU compendium [8]. This allowed for estimating the full economic cost of the time allocated to service contacts, time spent delivering the intervention at the patient level, and time spent for management and training activities at the aggregate level, as described above. For the feasibility-related objectives, we applied the same unit costs to HES and CSRI data.

**Table 2:** Unit costs.

| Service | £ sterling | Source |
| --- | --- | --- |
| **Hospital services** |  |  |
| Inpatient stay | 3478 | NHS Reference Cost 22/23 [23] |
| Outpatient appointment |  |  |
| Adult mental health service | 304 | NHS Reference Cost 22/23 |
| Audiology service | 120 | NHS Reference Cost 22/23 |
| Breast surgery service | 198 | NHS Reference Cost 22/23 |
| Cardiology service | 168 | NHS Reference Cost 22/23 |
| Cardiothoracic surgery service | 224 | NHS Reference Cost 22/23 |
| Clinical immunology and allergy service | 129 | NHS Reference Cost 22/23 |
| Clinical haematology service | 177 | NHS Reference Cost 22/23 |
| Clinical oncology service | 140 | NHS Reference Cost 22/23 |
| Dental medicine service | 217 | NHS Reference Cost 22/23 |
| Dermatology service | 162 | NHS Reference Cost 22/23 |
| Diabetes service | 197 | NHS Reference Cost 22/23 |
| Diagnostic imaging service | 55 | NHS Reference Cost 22/23 |
| Ear nose and throat service | 155 | NHS Reference Cost 22/23 |
| Emergency medicine service | 195 | NHS Reference Cost 22/23 |
| Endocrinology service | 193 | NHS Reference Cost 22/23 |
| Gastroenterology service | 158 | NHS Reference Cost 22/23 |
| General surgery service | 169 | NHS Reference Cost 22/23 |
| Genitourinary medicine service | 203 | NHS Reference Cost 22/23 |
| Gynaecology service | 191 | NHS Reference Cost 22/23 |
| Hepatology service | 195 | NHS Reference Cost 22/23 |
| Maxillofacial surgery service | 189 | NHS Reference Cost 22/23 |
| Neurology service | 220 | NHS Reference Cost 22/23 |
| Occupational therapy service | 122 | NHS Reference Cost 22/23 |
| Ophthalmology service | 136 | NHS Reference Cost 22/23 |
| Orthopaedic service | 160 | NHS Reference Cost 22/23 |
| Pain management service | 220 | NHS Reference Cost 22/23 |
| Physiotherapy service | 104 | NHS Reference Cost 22/23 |
| Podiatry service | 97 | NHS Reference Cost 22/23 |
| Rehabilitation medicine service | 248 | NHS Reference Cost 22/23 |
| Respiratory medicine service | 188 | NHS Reference Cost 22/23 |
| Rheumatology service | 173 | NHS Reference Cost 22/23 |
| Sleep Medicine service | 132 | NHS Reference Cost 22/23 |
| Speech and language therapy service | 190 | NHS Reference Cost 22/23 |
| Stroke medicine service | 303 | NHS Reference Cost 22/23 |
| Transient ischaemic attack service | 268 | NHS Reference Cost 22/23 |
| vascular surgery service | 191 | NHS Reference Cost 22/23 |
| Day patient visit | 894 | NHS Reference Cost 22/23 |
| A&E (walk in) | 136 | NHS Reference Cost 22/23 |
| A&E (admitted) | 455 | NHS Reference Cost 22/23 |
| **Other health and social services** |  |  |
| Intermediate care team | 12 | PSSRU P43 22/23 [17] |
| **Community contacts** |  |  |
| Physiotherapy at surgery (session) | 71 | NHS Reference Cost 22/23 |
| Physiotherapy home (session) | 123 | NHS Reference Cost 22/23 (1:0.73 Ratio of direct to indirect time on home visits) |
| Occupational therapy at surgery (session) | 56 | NHS Reference Cost 22/23 |
| Occupational therapy home (session) | 97 | NHS Reference Cost 22/23 |
| Speech and language therapy at surgery (session) | 112 | NHS Reference Cost 22/23 |
| Speech and language therapy home (session) | 211 | NHS Reference Cost 22/23 |
| **Other community contacts** |  |  |
| General practice (10 minutes) | 49 | PSSRU P63 22/23 |
| General practice (home visit) | 120 | PSSRU P167 10/11 [24] |
| General practice nurse (hour) | 53 | PSSRU P62 22/23 |
| General practice nurse (home visit) | 44 | PSSRU P167 10/11 |
| Therapy assistant at surgery / phone/ online (hour) | 36 | PSSRU P56 22/23 |
| Therapy assistant at home (hour) | 62 | PSSRU P153 10/11 |
| Health care support worker at surgery /phone (hour) | 15 | PSSRU P163 10/11 |
| Health care support worker at home (hour) | 24 | PSSRU P163 10/11 |
| Community Nurse, district nurse, health visitor (hour) | 35 | PSSRU P159 10/11 |
| Consultant (hour) | 143 | PSSRU P62 22/23 |
| Psychologist (hour) | 36 | PSSRU P166 10/11 |
| Chiropodist (hour) | 22 | PSSRU P154 10/11 |
| Chiropractor (hour) | 22 | Assumed same unit cost as chiropodist |
| Osteopath (hour) | 22 | Assumed same unit cost as chiropodist |
| Dentist (hour) | 116 | PSSRU P68 22/23 |
| Optician (Ophthalmic and Vision Science Service - visit) | 20 | NHS Reference Cost 22/23 |
| Complementary therapist (Acupuncture for Pain Management, Physiotherapy Service - visit) | 96 | NHS Reference Cost 22/23 |
| **General services** |  |  |
| Day hospital (attendance) | 104 | PSSRU P95 02/03 [25] |
| Social club (visit) | 17 | PSSRU P49 94/95 [26] |
| Lunch club | 17 | PSSRU P49 94/95 |
| Drop-in centre | 17 | PSSRU P49 94/95 |
| **Social services** |  |  |
| Meals on wheels (meal) | 6 | PSSRU P125 12/13 [27] |
| Frozen meals on wheels (meal) | 6 | PSSRU P125 12/13 |
| Home help for personal care (hour) | 27 | PSSRU P78 22/23 (home care worker) |
| Home help for household tasks (hour) | 27 | PSSRU P78 22/23 (home care worker) |
| Home help for shopping assistance (hour) | 27 | PSSRU P78 22/23 (home care worker) |
| Social worker – saw face to face (hour) | 53 | PSSRU P75 22/23 |
| Social worker – spoke by telephone (hour) | 53 | PSSRU P75 22/23 |
| Social services – day care centre (attendance) | 106 | PSSRU P33 22/23 |
| Stay in residential (day) | 237 | PSSRU P31 22/23 |
| Stay in nursing home (day) | 197 | PSSRU P7 22/23 |
| **Other** |  |  |
| Unpaid help from family and friends (hour) | 18 | ONS 2023 [28] |

### Outcome measures

The exploratory cost-effectiveness analysis was based on the primary outcome measure for the trial, the self-reported Nottingham Extended Activities of Daily Living Scale (NEADL [9]; for consistency, we followed the same calculation approach here as used for the analysis and reporting of the primary trial outcomes). The exploratory cost-utility analysis was based on Quality-Adjusted Life Years (QALYs) derived from utility scores applied to health-related quality of life descriptions obtained using the EQ-5D-5L instrument [10]. Both measures were completed by the participants or their principal carer at baseline, 6-months, 12-months, and 24-months post randomisation. UK utility values for the EQ-5D-5L were derived using the approach recommended by the National Institute for Health & Care Excellence (NICE), which is currently using the validated mapping function from the existing EQ-5D-3L[11]. These were then used to form QALYs over the 6-month, 12-month, and 24-month periods using area-under-the-curve methods assuming a linear change between any two adjacent time points [12].

### Analyses

Analyses were carried out on participant-level clinical trial data and according to the ITT principle [13]. All data relevant for the economic evaluation, including data collected from the CSRI, EQ-5D-5L and NEADL, were checked and cleaned. Mean imputation was used to fill in each missing EQ-5D-5L index value at baseline independent of the treatment arm. The same approach was used to impute missing data for NEADL and the different categories of costs at baseline. For missing EQ-5D-5L index values and NEADL scores at all follow-up points, except 24-months (see below for explanation), we used the diagnostic procedure outlined in Faria et al.’s guide [14] for missing data imputations for economic evaluations. The data were assumed missing at random (MAR), and the missing observations were replaced with a set of imputed values through multiple imputations drawn from posterior predictive distribution given the missing observed data. Missing cost data at all follow up points, except 24-months, were imputed at the different categories of costs using the same approach outlined for dealing with missing EQ-5D-5L index values at follow up. Attempts to impute missing data at 24-months produced an error (“VCE is not positive definite”, Stata version 18). This error was expected considering the substantially low rate of completed CSRI questionnaires at 24-months (due to the curtailing of follow-up for some participants). which produced insufficient observations. Therefore, the small sample size at 24-months resulted in the imputation model struggling to produce a positive definite variance-covariance matrix of estimators. All analyses were completed using STATA version 18.

Resource use is reported descriptively, using the mean and standard deviation, and not compared statistically principally to avoid problems associated with multiple testing but also due to the exploratory nature of the economic evaluation and the focus on exploring estimates from different data/analytical approaches.

Differences in mean total costs with associated bias corrected 95% confidence intervals between the two arms from NHS/PSS and from societal perspectives over the period from baseline to 12-month were analysed using bootstrap multi-level mixed effect linear models adjusting for participants’ baseline costs with randomisation stratification variables as covariates.

The mean difference in QALYs over the period from baseline to 12-month period was estimated using bootstrap multi-level mixed effect model with associated bias corrected 95% confidence intervals. Baseline utility score with randomisation stratification variables were used as covariates. The mean difference in NEADL was estimated using the model specification mentioned for estimating the difference in QALYs.

Probabilities that the true (unobserved) population difference in mean cost, QALYs and NEADL between the two treatment arms are positive (i.e., cost, QALYs, NEADL score are higher in the intervention arm when compared to control) based on trial sample information were estimated.

To make exploratory inferences about the cost-effectiveness of the intervention, a joint distribution of mean difference in cost and QALYs/NEADL was produced by fitting cost and outcome models jointly to each bootstrap replicate. Results are presented as scatterplots of these distributions.

All costs are reported at 2022/23 levels using the Personal Social Services Pay & Price Index and the NHS Cost Inflation Index [15, 16, 17] to adjust unit costs as necessary. Discounting to adjust for the differential timing of costs and quality-adjusted life years (QALYs) estimated between 12– and 24-month period was implemented using 3.5% discount rate [18].

## RESULTS

### Participants and follow-up

A total of 5984 participants were screened: 1029 (17.2% of screened) were approached, 969 (94.2% of approached, 16.2% of screened) were eligible and 334 (34.6% of those eligible; 5.6% of those screened) were registered into the trial; 181 participants were registered to the GSG arm and 153 to the usual care arm. Participants across the treatment arms were similar in terms of age (average of 69 years, range 23-98), gender (60.4% female), living arrangements, disability, stroke severity and cognitive impairment. However, there were some differences in ethnicity of participants; 94.4% were White British in the GSG arm compared to 85.4% in the usual care arm – possibly due to regional variations between the recruiting sites. There were also differences in the type and presentation of stroke. The percentage of participants with primary intracerebral haemorrhage was 15.0% in the GSG arm and 11.3% in the UC arm. The percentage of participants with left hemiparesis was 51.7% in the GSG arm and 36.4% in the UC arm. Details of withdrawals are provided elsewhere but baseline characteristics were very similar between those who did and did not withdraw.

Follow-up rates were similar across the two arms. Baseline characteristics were broadly similar across those included in the primary analysis compared to those lost to follow-up. However, there were some small differences; participants lost to follow-up were slightly more likely to live alone (33.1% vs 27.0% of those followed up) or have speech and language impairments (13.1% vs 7.9% of those followed up).

### Intervention delivery

For the 181 participants receiving the GSG intervention, 103 (57.8%) inpatient therapy records, 51 (28.3%) nursing records, and 76 (42.8%) community records were returned by sites.

From the inpatient records, 74.8% (77/103) of participants were provided the GSG guide, and for 71.8% (74/103) of participants staff shared their prescription with the nursing team and used the checklist to discuss the guide with the participant. Weekly nursing records were completed for a median of 1 week per participant, ranging up to 5 weeks. Daily tasks such as prompting standing and moving according to prescription and encouraging participant to use the guide to record standing and moving were completed on approximately 90% of days (ranging from 89.3% to 91.8% of days across tasks).

From the community records, 86.8% (66/76) of participants had a GSG guide, either from hospital or replaced by community staff, however only 14.5% (11/76) participants had the optional prompt magnet provided. Weekly community records were completed for a median of 4 weeks per participant, ranging up to 11 weeks and including a total of 380 days across the 76 participants. Daily tasks of checking the traffic light status and prescription and prompting participant to stand and move and use the guide were completed for approximately 87% of days (ranging from 85.8% to 91.8% of days across tasks).

### Resource use

Resource use at baseline (for the previous 3 months), 6 months (for the previous 6 months), 12 months (for the previous 6 months), and 24 months (for the previous 6 months) is detailed in Tables S1, S2, S3 and S4 respectively. The pattern of use of resources at baseline is broadly similar in both arms but with some observed differences in home-based services and unpaid help; the GSG arm received considerably more unpaid help from people living with the participants (23.66 vs. 4.12 hours on average), with a high variability (SD=146.46 vs. 33.14). At the follow-up points, the use of hospital services is observably higher in the GSG arm compared to the usual care arm. For example, at 6 months follow-up, the average number of inpatient nights was 3.01 in the GSG arm compared to 1.47 in the usual care arm, and the number of outpatient appointments was 3.20 in the GSG arm compared to 2.09 in the usual care arm. The large standard deviations suggest that there are a few participants who had more frequent inpatient admissions. Use of community services were similar between randomised groups. However, the GSG arm relied more on home-based services and there were clear differences in the amount of unpaid help from carers living with participants, with higher (146.54 vs. 64.29 hours) average hours in the GSG arm compared to the usual care arm, though similarly the large standard deviation should be noted. The resource use data for 12 and 24 months shows that there are very few differences in use of services between randomised groups, with unpaid help significantly higher in the GSG arm at 12 months suggesting a greater reliance on informal caregiving.

### Costs

Total costs were higher in the GSG arm across baseline and all follow-ups, as detailed in Table 3, and Supplementary Appendix, Tables S5, S6, S7 and S8. At baseline, total NHS/PSS costs (£658 vs. £487) and societal costs (£2710 vs. £1452) were higher in the GSG arm mainly driven by higher costs for day patient care, A&E visits and unpaid help. At 6 and 12 months, the observed differences in NHS/PSS and societal costs between the two treatment arms were similar to baseline with the GSG arm costing more. The GSG arm showed higher variability which means there was wider cost dispersion and implying a highly skewed cost distribution with a standard deviation twice as the mean at 12 months follow up. The data points at 24 months were scarce making it difficult to report a meaningful observation.

**Table 3:** Adjusted mean difference in total costs, QALYs, and NEADL from NHS/PSS and societal perspectives, baseline to 12 months.

|  | GSG (Mean<sup>3</sup>) | Usual care (Mean) | Difference in means<sup>4</sup> | 95% CI |
| --- | --- | --- | --- | --- |
| Total cost, NHS/PSS<sup>1</sup> | 1091 | 865 | £846 | 140 to 1552 |
| Total cost, societal<sup>2</sup> | 11140 | 8989 | £5899 | -1397 to 13195 |
| QALYs | 0.524 | 0.657 | -0.062 | -0.106 to -0.018 |
| NEADL | 43.28 | 47.41 | -3.14 | -6.87 to 0.58 |

<sup>1</sup>Total NHS/PSS cost consists of total hospital services cost plus therapy services in the community, NHS community-based services and intervention costs; sum of costs assessed at 6m and 12m follow-ups.
<sup>2</sup>Total Societal cost consists of total NHS/PSS cost plus non-NHS community-based services, unpaid help and intervention costs; sum of costs assessed at 6m and 12m follow-ups.
<sup>3</sup>Calculated using data from complete case sample
<sup>4</sup>Calculated using bootstrap multi-level mixed effect linear models after multiple imputation of missing data adjusting for cluster-level randomisation stratification factors, baseline outcome score, type of stroke and time since stroke

### Outcomes

The mean NEADL score pre-stroke was 57.4, with slightly greater independence in the usual care arm (58.9 compared to 56.1). At 6– and 12-months after stroke, the mean NEADL score was approximately 45.3 with a median score of 50.0 and a wide interquartile range from 31 to 61 points. Mean NEADL score at 12 months was 43.3 (18.87) in the GSG arm vs 47.4 (17.11) in the usual care arm.

Average utility score by randomised group is detailed in Table 4. At baseline, and at all follow-up periods, although the differences were small, they result in lower QALYs over follow-up in the GSG arm compared to the usual care arm.

**Table 4:** EQ-5D-5L utility scores at baseline and all follow-up points.

| Assessment | GSG (N=180) |  |  | Usual care (N=151) |  |  | Overall (N=331) |  |  |
| --- | --- | --- | --- | --- | --- | --- | --- | --- | --- |
|  | (N) | (Mean) | (SD) | (N) | (Mean) | (SD) | (N) | (Mean) | (SD) |
| Baseline | 169 | 0.584 | 0.284 | 129 | 0.679 | 0.260 | 298 | 0.625 | 0.277 |
| 6-month | 87 | 0.531 | 0.347 | 93 | 0.638 | 0.293 | 180 | 0.586 | 0.323 |
| 12-month | 101 | 0.480 | 0.372 | 95 | 0.635 | 0.312 | 196 | 0.555 | 0.352 |
| 24-month | 42 | 0.338 | 0.402 | 32 | 0.604 | 0.372 | 74 | 0.453 | 0.409 |

### Exploratory economic evaluation

Total costs were higher in the GSG arm across baseline and all follow-ups (Supplementary Appendix, Tables S5, S6, S7 and S8). This is likely due to the GSG arm having higher estimates of unpaid care at baseline which remained evident at follow-ups, coupled with higher use of hospital services (Supplementary Appendix, Tables S1, S2 S3 and S4). Utility estimates also suggested lower QALYs for the GSG arm over the follow-up period (Table 3). In combination, from both NHS/PSS and societal perspectives, costs are higher in the GSG arm, whilst outcomes measured in QALYs are worse, suggesting that the GSG intervention is unlikely to be cost-effective. This is further evidenced by the bootstrapped replications in **Error! Reference source not found.** and **Error! Reference source not found.** which show that almost all replications lie in the northwest quadrant of the cost-effectiveness plane (which indicates that the GSG arm is more costly and less effective than the usual care arm, meaning they are dominated and not cost-effective).

### Feasibility objectives

HES data suggested that hospital service use is consistently higher across both randomised groups compared to the self-reported CSRI data, resulting in higher estimates for hospital costs (Table 5). Of note, estimates derived from HES did not provide evidence of higher costs in the GSG arm compared to the usual care arm. Therefore, the direction of difference in service use and costs, which for self-report were consistently higher in the GSG arm compared to the usual care arm, was not mirrored in the equivalent estimates derived from HES data.

**Table 5:** HES data: mean (SD) resource use and costs over the follow-up period.

|  | GSG (n=181) |  | Usual care (n=153) |  | Difference |
| --- | --- | --- | --- | --- | --- |
|  | Mean | SD | Mean | SD |  |
| **Service use** |  |  |  |  |  |
| Outpatient appointments 0-12 months | 6.96 | 8.86 | 7.71 | 9.66 | -0.75 |
| Outpatient appointments 12-24 months | 4.37 | 12.06 | 6.17 | 12.93 | -1.8 |
| Daycase 0-12 months | 0.67 | 1.57 | 0.64 | 1.09 | 0.03 |
| Daycase 12-24 months | 0.86 | 4.55 | 0.65 | 1.28 | 0.21 |
| Inpatient nights 0-12 months | 11.57 | 21.92 | 8.24 | 14.62 | 3.33 |
| Inpatient nights 12-24 months | 6.45 | 23.66 | 4.42 | 15.74 | 2.03 |
| **Costs** |  |  |  |  |  |
| Outpatient total cost 0-12 months | 1449 | 1853 | 1595 | 2050 | -147 |
| Outpatient total cost 12-24 months | 917 | 2531 | 1278 | 2710 | -361 |
| Daycase total costs 0-12 months | 598 | 1404 | 573 | 971 | 25 |
| Daycase total costs 12-24 months | 766 | 4064 | 578 | 1144 | 187 |
| Inpatient total cost 0-12 months | 2210 | 1679 | 2432 | 1600 | -223 |
| Inpatient total cost 12-24 months | 788 | 1460 | 1205 | 1660 | -417 |

## DISCUSSION

This pilot trial explored the potential cost-effectiveness of Get Set Go (GSG), a complex multi-sector intervention to reduce sedentary behaviour among stroke survivors. It also explored the potential impacts of using alternative data sources for such an evaluation, with a view to informing the design of a future definitive trial focusing on a similar intervention and/or similar study population.

The exploratory cost-effectiveness analysis indicated that the GSC intervention is unlikely to be cost-effective compared to usual care among the examined patient group. In the base case findings (which relied on self-reported resource use data), at 6 months, the GSG arm showed slightly higher hospital service use and costs compared to the usual care group, alongside lower EQ-5D-5L utility scores and thus QALYs. Importantly, despite randomisation, the GSG arm had higher service use at baseline and this pattern persisted at each follow-up. The cost-effectiveness analyses thus indicated that the GSG intervention was not cost-effective compared to usual care, which was supported by evaluation of statistical uncertainty.

These analyses carried various strengths and weaknesses. In terms of the overall trial, COVID-19 caused issues in delivery, which led to slower than predicted recruitment and underpowering to detect statistical significance. Given the circumstances, included sites likely represented stroke services with a higher-than-average interest and willingness to changing standing and moving practice. The population was less severe than expected, which may have been due to recruitment bias. There was also an imbalance between arms in some participant characteristics, potentially due to case mix and regional population variation between sites rather than selection bias. Follow-up rates were lower than anticipated and affected running the analyses over a 24-month period. There was also a high level of missing data on intervention delivery. More specifically with regards to the economic evaluation, despite its exploratory nature, we undertook a comprehensive approach based on a robust and standard framework and relevant probabilistic sensitivity analyses to explore the uncertainty in the data. The longitudinal perspective that the 24-month follow-up provides is longer than many trials in this patient population, although it comes with its challenges as evidenced by the low follow-up rates at the final assessment.

Our finding that secondary care was the second largest driver (after unpaid care) of estimated total societal costs for the exploratory cost-effectiveness analysis highlights the pertinence of examining the relative merits of self-reported data versus routine care records for the purposes of such an evaluation. However, the comparison of secondary care use and costs based on HES data rather than retrospective self-reported accounts revealed some discrepancies which point to a potential recall bias in self-reported data. These in turn indicate uncertainty over the strength of the exploratory cost-effectiveness findings (over and above those related to the pilot nature of the trial).

It is overall challenging to draw direct comparisons of estimates generated by the two data sources since the self-reported data related to three 6-month periods over the two years of follow-up, leaving a gap in observation between 12 and 18 months, whereas the HES data covered the whole period of follow-up. The gap in the self-reported data was a necessary limitation since the trial did not incorporate an 18-month follow-up assessment and, at 24 months, it was not considered reliable to ask about a retrospective period of greater than 6 months due to the increased risk of recall bias [19]. However, we find that service use is clearly higher when estimated from the HES data compared to self-reported data, leading to higher total cost estimates for the sample based on HES data compared to self-reported data. Given that a different pattern of hospital service use is found in the HES data, the cost-effectiveness findings are likely to be different, perhaps reflecting more uncertainty in the difference in cost, though the observed quality of life outcomes would remain worse in the GSG arm compared with the usual care arm.

We observed a low level of agreement between self-report and HES data, with self-reported data consistently underestimating the number of contacts with secondary care services. Given the contribution of these resource items to total health/PSS costs, it raises questions about the reliability of using self-reported accounts of secondary care use for evaluations in this context.

The 24-month data demonstrate that, in this patient group, hospital services continue to be used longer term and costs remain high. This highlights the importance of collecting longer term data in this group for the purposes of generating reliable estimates of cost-effectiveness of interventions targeted at stroke survivors. Given the findings discussed above related to the advantages of using HES data over self-reported data, and the low rates of follow-up at 24 months, HES is likely to be a useful facilitator of longer-term follow-up for this patient group. HES data are also likely to be a reliable and convenient replacement for self-report service use for participants for whom data are missing, and indeed for secondary care service use for all participants.

The breadth of the cost assessment was a particular strength. Stroke survivors are known to experience a vast range of ongoing physical and mental health consequences long after their stroke and a reliable assessment of cost-effectiveness of any intervention for this patient population needs to be inclusive of a wide range of potential impacts and associated care costs. This is a key rationale for employing a self-reported mode of data collection since this is the only way to assess the diverse range of health, social care and societal costs in the absence of routine data sources for all such impacts. Health care records are understood to be reliable for the purposes of care planning and research, but there remain challenges in accessing secondary care data in a timely way, and in accessing linked primary and secondary care data for the purposes of a multi-centre evaluation context. Meanwhile, routine data for other community-based care data remain challenging to collect and access due to the breadth of agencies involved in providing such care. Further, where there is an interest in understanding the impacts on unpaid carers, patients and their (possibly multiple) unpaid helpers remain the most appropriate informants given the complexity and nuance of collecting such evidence. Therefore, self-reported data collection for resource use for this patient group has long remained an important source of data to enable comprehensive economic evaluation. However, as evidenced in this study, self-reported data are also associated with a high risk of missing data due to loss of follow-up, which in turn impacts the reliability of an evaluation.

Given the complexity of evaluating complex interventions among stroke survivors, several key lessons emerge from this study. First, the need for longer-term follow-up is evidenced by the significant care costs that are sustained over 24 months. Second, the importance of capturing unpaid care is evident by the costs for this overshadowing formal care costs. Third, given the discrepancies between self-reported and routine health care resource use data – but coupled with the importance of maintaining broad evaluative perspectives for this patient population – it is likely that a hybrid approach to data collection would be appropriate. Routine records would help capture key secondary care costs as accurately as possible, while supplementary self-reported data collection for other important resource use such as unpaid care and rehabilitation therapies would ensure relevant impacts are accounted for, but with reduced data collection burdens placed upon participants. This is consistent with recent recommendations by Franklin & Thorn [20] who reviewed the use of resource use estimates obtained from self-reported methods and routine records and found that while the latter may overall be considered preferable for accuracy, remaining logistical challenges for (related to data governance and access) and the inability of such data to support a societal perspective in an economic evaluation, preclude routine records from being the mainstay of resource use data collection. However, questionnaires developed for the purposes of self-reported resource use data collection are likely to require careful consideration of various factors within the evaluation context [21, 22] and possibly greater standardisation [23]. Finally, proportionate, hybrid approaches to estimating the costs of complex interventions such as GSG are likely to be appropriate in a multi-centre context (compared with patient-level micro-costing approaches) given their relatively small size in the context of overall health/PSS and societal costs.

## Data Availability

As this was a feasibility trial to inform a definitive trial, sharing of the trial data set is not anticipated; however, any data requests should be sent to the corresponding author and would be subject to review by a sub-group of the trial team which will include the data guarantor, Professor Farrin. All data sharing activities would require a data sharing agreement.

## CONCLUSIONS

This exploratory analysis indicates that GSG is unlikely to be cost-effective. It also highlights important methodological lessons for future similar trials, especially for similar patient populations. Routine data, such as HES, may provide a more reliable and less burdensome source for estimating secondary care costs, whereas self-report remains important for capturing unpaid care and community-based resource use. A hybrid data collection approach is recommended for future economic evaluations in stroke rehabilitation.

## Study registration

RECREATE Programme Workstream 5: ISRCTN registry, ISRCTN82280581. Registered on 1 April 2020.

## Ethics

Ethics approval for the study was granted by the Yorkshire and The Humber – Bradford-Leeds Research Ethics Committee (19/YH/0403).

## Acknowledgements

We thank the huge efforts of many who helped delivered this study (including a number of services which progressed to set-up the trial prior to the pandemic but were then unable to continue post COVID-19). Particular thanks go to all the clinical teams who participated, it is hugely appreciated that they took on this additional research-related work at a time when they were also addressing the multiple challenges of the COVID-19 pandemic. They supported the research team in adapting the research, for example facilitating on-line training rather than in-person. We would like to acknowledge those stroke services that have not only supported this programme of work but our previous research over many years. Thank you to all the survivors of stroke and their families who participated in this research and those who contributed through engagement in discussions at local groups and ad hoc conversations over the years, their input was invaluable. Thank you to our sponsor Bradford Teaching Hospitals NHS Foundation Trust (BTHFT) and specifically Jane Dennison, who supported us through the COVID-19-related challenges. We are extremely grateful for inputs from the members of the Programme Steering Committee (PSC) who have supported the research team throughout implementation of the RECREATE programme: Dawn Skelton (Chair), Jacqui Morris, Jonathan Mant, Mark Kelson, Saima Siddiqui (NIHR representative) and Amanda North (stroke survivor). We would like to thank colleagues in the Academic Unit of Ageing and Stroke Research (BTHFT and University of Leeds) and the Clinical Trials Research Unit (University of Leeds) who provided support and advice during the study.

---

**Figure 1:**
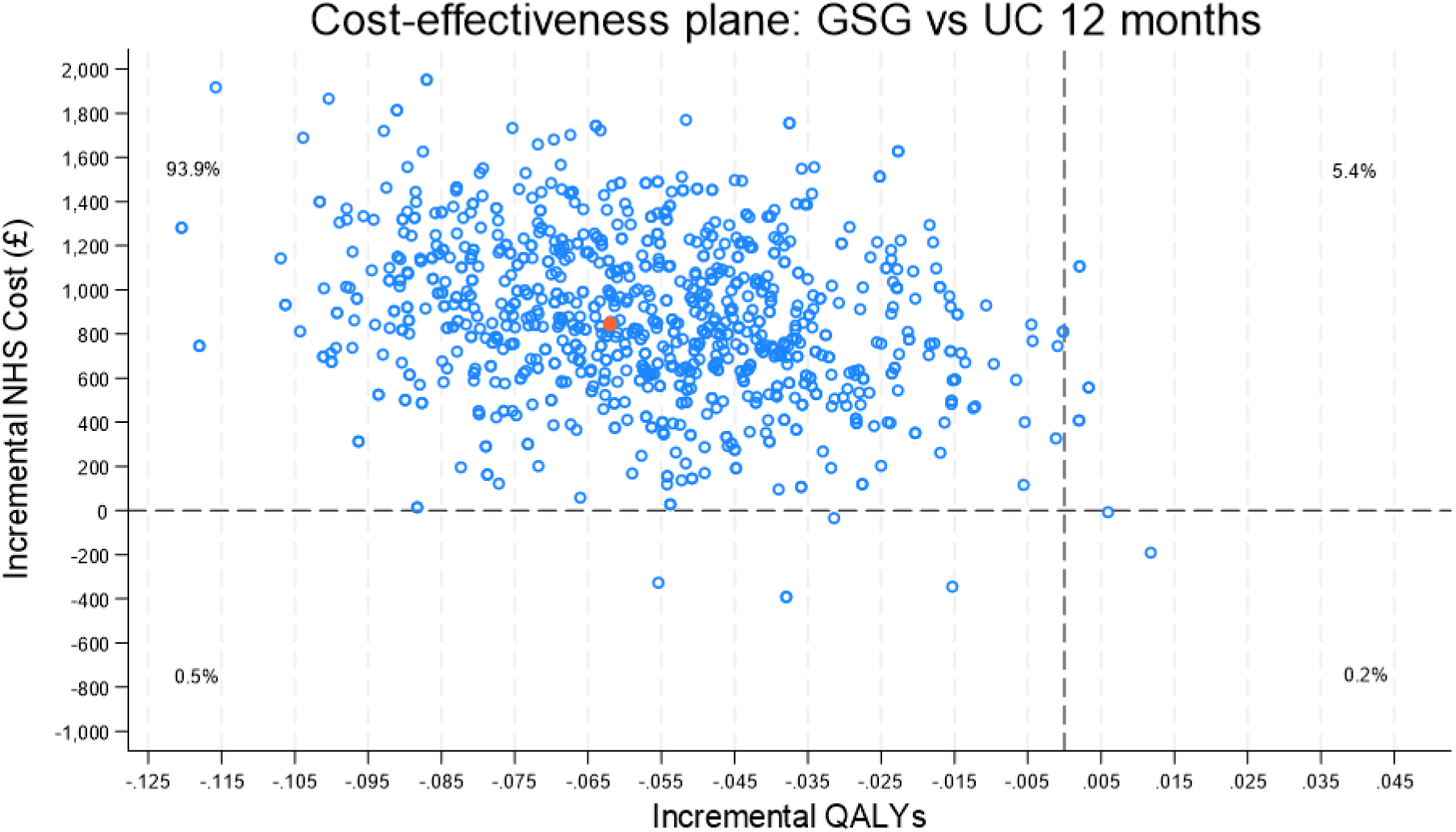
Cost-effectiveness plane for costs from NHS & PSS perspective and QALY differences.

**Figure 2:**
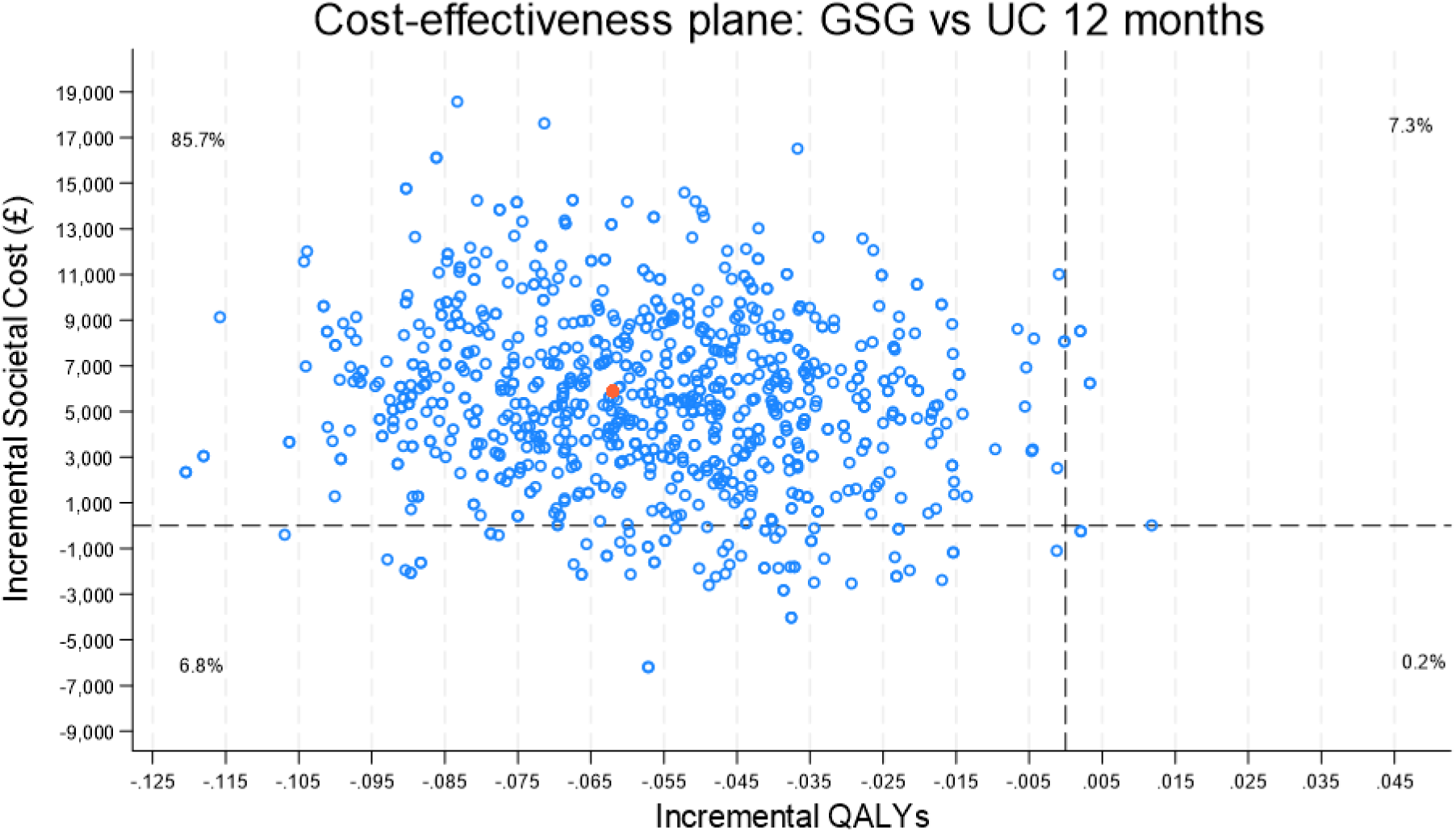
Cost-effectiveness plane for costs from societal perspective and QALY differences.

## Supplementary Appendix

**Box S1.**
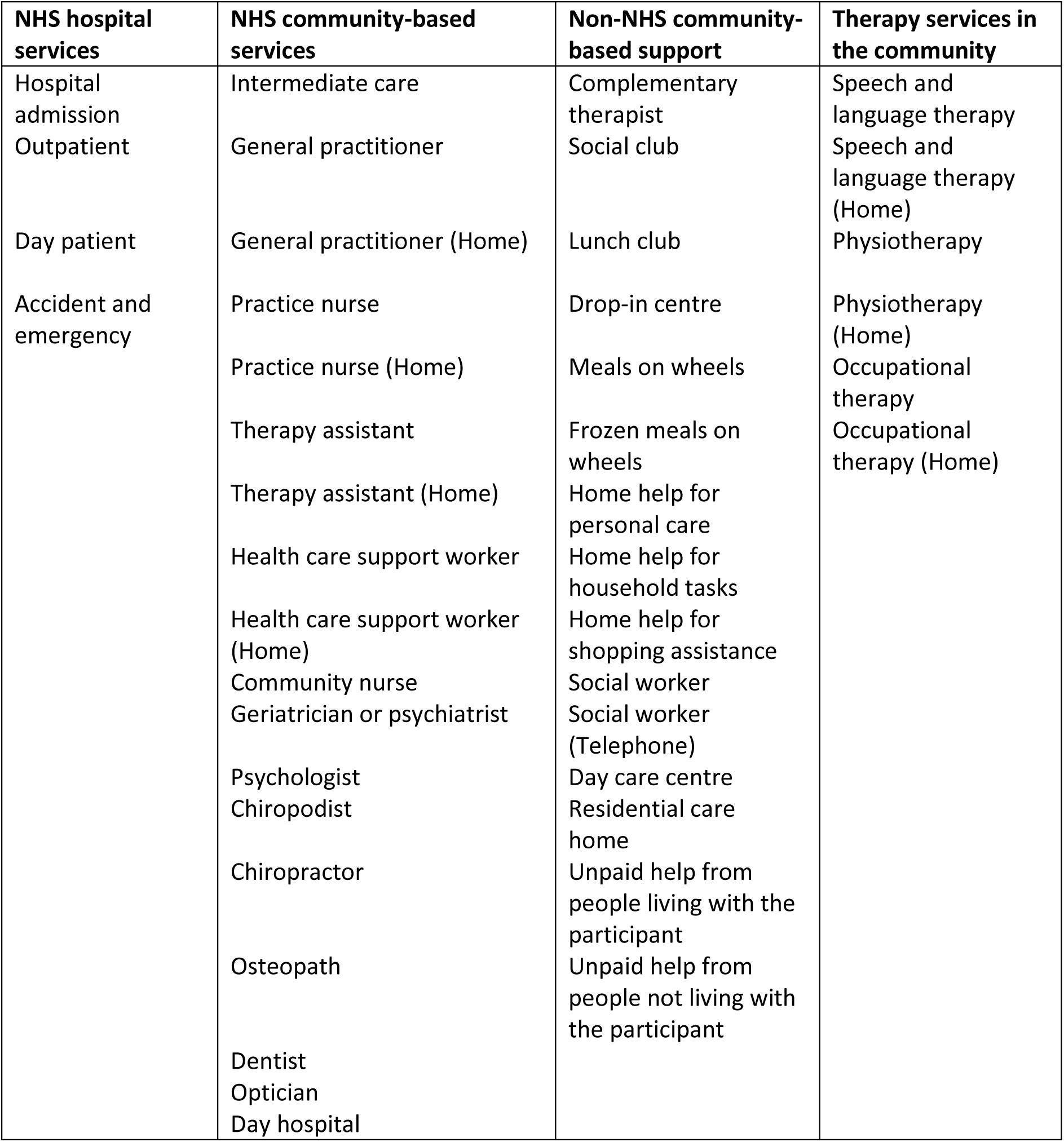
Identified resources for measurement.

**Table S1:** Resource use: number of participants using a service and mean (SD) contacts at baseline (for the previous 3 months)

|  | GSG (N=180) |  |  |  | Usual care (N=151) |  |  |  | Overall (N=331) |  |  |  |
| --- | --- | --- | --- | --- | --- | --- | --- | --- | --- | --- | --- | --- |
| Resource | (N) returned | (N) used | (Mean) | (SD) | (N) returned | (N) used | (Mean) | (SD) | (N) returned | (N) used | (Mean) | (SD) |
| **Hospital services** |  |  |  |  |  |  |  |  |  |  |  |  |
| Hospital admission | 169 | 10 | 0.34 | 2.02 | 150 | 8 | 0.28 | 1.52 | 319 | 18 | 0.31 | 1.80 |
| Outpatient | 167 | 49 | 0.63 | 1.32 | 149 | 38 | 0.58 | 2.45 | 316 | 87 | 0.61 | 1.94 |
| Day patient | 166 | 9.00 | 0.06 | 0.29 | 145 | 1.00 | 0.01 | 0.08 | 311 | 10 | 0.04 | 0.22 |
| Accident and emergency | 169 | 20.00 | 0.12 | 0.32 | 147 | 15.00 | 0.10 | 0.30 | 316 | 35 | 0.11 | 0.31 |
| **NHS community-based services** |  |  |  |  |  |  |  |  |  |  |  |  |
| Intermediate care | 156 | 13.00 | 0.09 | 0.39 | 127 | 1.00 | 0.08 | 0.89 | 283 | 14 | 0.09 | 0.66 |
| General practitioner | 171 | 91.00 | 1.27 | 2.06 | 151 | 74.00 | 1.13 | 2.24 | 322 | 165 | 1.21 | 2.14 |
| General practitioner (Home) | 167 | 8.00 | 0.05 | 0.29 | 150 | 4.00 | 0.02 | 0.14 | 317 | 12 | 0.04 | 0.23 |
| Practice nurse | 167 | 53.00 | 0.55 | 1.33 | 150 | 24.00 | 0.28 | 1.18 | 317 | 77 | 0.42 | 1.27 |
| Practice nurse (Home) | 166 | 4.00 | 0.01 | 0.11 | 149 | 1.00 | 0.01 | 0.16 | 315 | 5 | 0.01 | 0.14 |
| Therapy assistant | 166 | 4.00 | 0.01 | 0.11 | 150 | 2.00 | 0.01 | 0.08 | 316 | 6 | 0.01 | 0.10 |
| Therapy assistant (Home) | 169 | 3.00 | 0.01 | 0.15 | 149 | 2.00 | 0.03 | 0.26 | 318 | 5 | 0.02 | 0.21 |
| Health care support worker | 168 | 3.00 | 0.03 | 0.39 | 148 | 1.00 | 0.07 | 0.82 | 316 | 4 | 0.05 | 0.63 |
| Health care support worker (Home) | 166 | 3.00 | 0.11 | 1.41 | 149 | 2.00 | 0.08 | 0.83 | 315 | 5 | 0.10 | 1.17 |
| Community nurse | 168 | 10.00 | 0.08 | 0.46 | 150 | 4.00 | 0.08 | 0.61 | 318 | 14 | 0.08 | 0.54 |
| Geriatrician or psychiatrist | 169 | 1.00 | 0.00 | 0.00 | 150 | 0.00 | 0.00 | 0.00 | 319 | 1 | 0.00 | 0.00 |
| Psychologist | 169 | 2.00 | 0.01 | 0.15 | 149 | 1.00 | 0.02 | 0.25 | 318 | 3 | 0.02 | 0.20 |
| Chiropodist | 169 | 18.00 | 0.14 | 0.85 | 147 | 11.00 | 0.08 | 0.32 | 316 | 29 | 0.11 | 0.65 |
| Chiropractor | 169 | 5.00 | 0.02 | 0.19 | 147 | 2.00 | 0.02 | 0.25 | 316 | 7 | 0.02 | 0.22 |
| Osteopath | 168 | 2.00 | 0.00 | 0.00 | 150 | 3.00 | 0.02 | 0.14 | 318 | 5 | 0.01 | 0.10 |
| Dentist | 171 | 34.00 | 0.23 | 0.66 | 150 | 39.00 | 0.30 | 0.64 | 321 | 73 | 0.27 | 0.65 |
| Optician | 167 | 31.00 | 0.23 | 0.57 | 147 | 26.00 | 0.15 | 0.43 | 314 | 57 | 0.19 | 0.51 |
| Complementary therapist | 169 | 2.00 | 0.07 | 0.93 | 146 | 1.00 | 0.02 | 0.25 | 315 | 3 | 0.05 | 0.70 |
| Day hospital | 169 | 2.00 | 0.02 | 0.23 | 149 | 2.00 | 0.01 | 0.08 | 318 | 4 | 0.01 | 0.18 |
| **Non-NHS community-based services** |  |  |  |  |  |  |  |  |  |  |  |  |
| Social club | 169 | 8.00 | 0.17 | 1.34 | 147 | 3.00 | 0.20 | 1.45 | 316 | 11 | 0.18 | 1.39 |
| Lunch club | 169 | 4.00 | 0.11 | 0.98 | 148 | 1.00 | 0.08 | 0.99 | 317 | 5 | 0.09 | 0.98 |
| Drop-in centre | 162 | 1.00 | 0.00 | 0.00 | 142 | 1.00 | 0.08 | 1.01 | 304 | 2 | 0.04 | 0.69 |
| Meals on wheels | 170 | 3.00 | 0.01 | 0.08 | 151 | 1.00 | 0.08 | 0.98 | 321 | 4 | 0.04 | 0.67 |
| Frozen meals on wheels | 169 | 4.00 | 0.02 | 0.23 | 151 | 1.00 | 0.00 | 0.00 | 320 | 5 | 0.01 | 0.17 |
| Home help for personal care | 167 | 5.00 | 0.15 | 1.88 | 151 | 0.00 | 0.00 | 0.00 | 318 | 5 | 0.08 | 1.35 |
| Home help for household tasks | 166 | 10.00 | 0.42 | 2.52 | 150 | 4.00 | 0.30 | 2.72 | 316 | 14 | 0.36 | 2.61 |
| Home help for shopping assistance | 164 | 5.00 | 0.15 | 1.90 | 150 | 1.00 | 0.00 | 0.00 | 314 | 6 | 0.08 | 1.37 |
| Social worker | 166 | 2.00 | 0.00 | 0.00 | 147 | 0.00 | 0.00 | 0.00 | 313 | 2 | 0.00 | 0.00 |
| Social worker (Telephone) | 167 | 3.00 | 0.00 | 0.00 | 148 | 0.00 | 0.00 | 0.00 | 315 | 3 | 0.00 | 0.00 |
| Day care centre | 167 | 3.00 | 0.00 | 0.00 | 150 | 0.00 | 0.00 | 0.00 | 317 | 3 | 0.00 | 0.00 |
| Residential care home | 163 | 1.00 | 0.52 | 6.58 | 146 | 0.00 | 0.00 | 0.00 | 309 | 1 | 0.27 | 4.78 |
| **Therapy services in the community** |  |  |  |  |  |  |  |  |  |  |  |  |
| Speech and language therapy | 166 | 0.00 | 0.00 | 0.00 | 149 | 0.00 | 0.00 | 0.00 | 315 | 0 | 0.00 | 0.00 |
| Speech and language therapy (Home) | 163 | 0.00 | 0.00 | 0.00 | 146 | 1.00 | 0.00 | 0.00 | 309 | 1 | 0.00 | 0.00 |
| Physiotherapy | 171 | 6.00 | 0.10 | 0.59 | 151 | 10.00 | 0.14 | 0.75 | 322 | 16 | 0.12 | 0.67 |
| Physiotherapy (Home) | 168 | 2.00 | 0.01 | 0.15 | 150 | 4.00 | 0.09 | 0.86 | 318 | 6 | 0.05 | 0.60 |
| Occupational therapy | 169 | 2.00 | 0.02 | 0.24 | 148 | 0.00 | 0.00 | 0.00 | 317 | 2 | 0.01 | 0.18 |
| Occupational therapy (Home) | 166 | 2.00 | 0.01 | 0.16 | 147 | 1.00 | 0.00 | 0.00 | 313 | 3 | 0.01 | 0.11 |
| **Unpaid help** |  |  |  |  |  |  |  |  |  |  |  |  |
| Help from people living with participants | 171 | 21.00 | 23.66 | 146.46 | 150 | 9.00 | 4.12 | 33.14 | 321 | 30 | 14.52 | 109.51 |
| Help from people not living with participants | 163 | 8.00 | 4.98 | 41.31 | 148 | 5.00 | 3.29 | 25.43 | 311 | 13 | 4.17 | 34.59 |

**Table S2:**
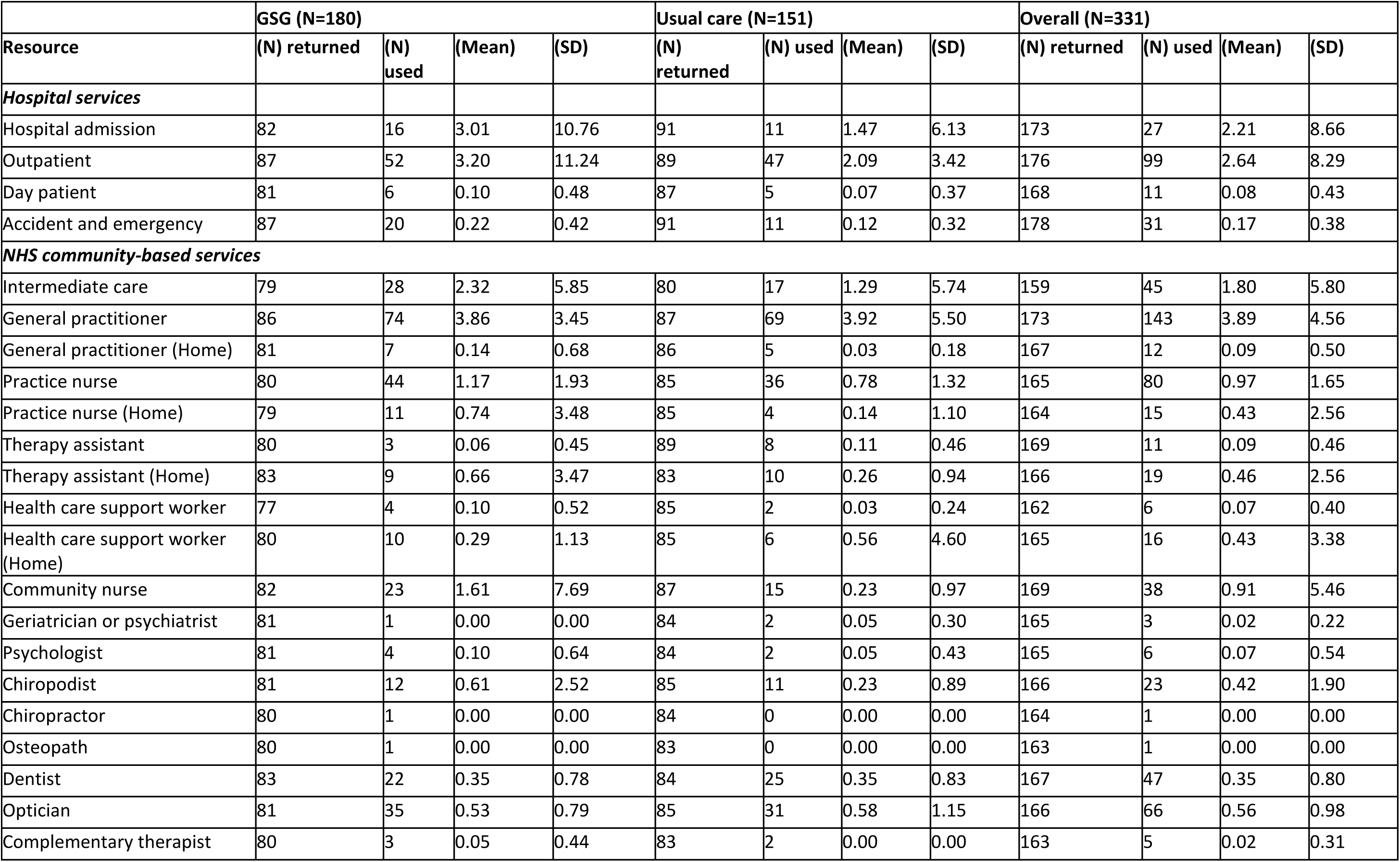
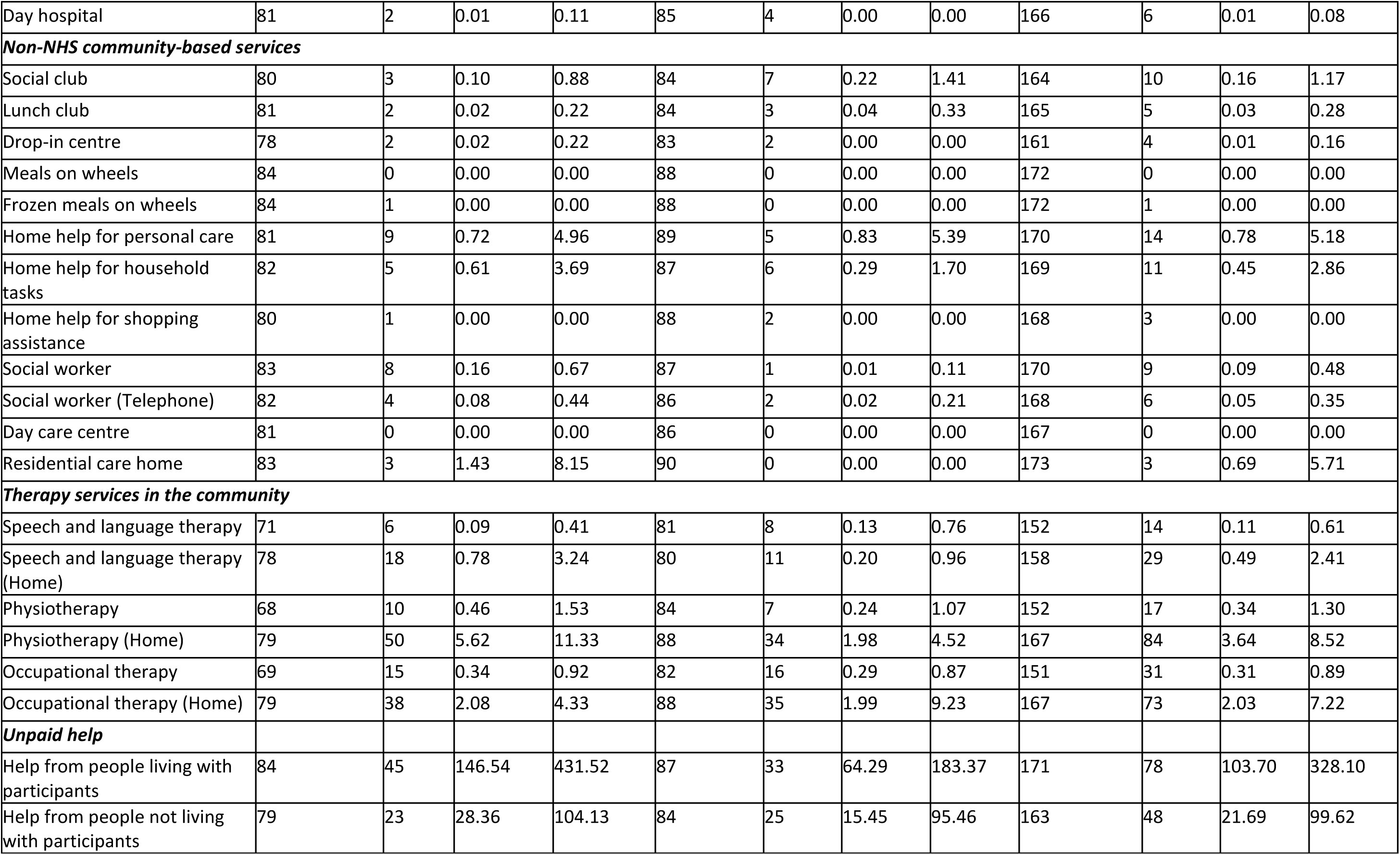
Resource use: number of participants using a service and mean (SD) contacts at 6-months (for the previous 6 months)

|  | GSG (N=180) |  |  |  | Usual care (N=151) |  |  |  | Overall (N=331) |  |  |  |
| --- | --- | --- | --- | --- | --- | --- | --- | --- | --- | --- | --- | --- |
| Resource | (N) returned | (N) used | (Mean) | (SD) | (N) returned | (N) used | (Mean) | (SD) | (N) returned | (N) used | (Mean) | (SD) |
| **Hospital services** |  |  |  |  |  |  |  |  |  |  |  |  |
| Hospital admission | 82 | 16 | 3.01 | 10.76 | 91 | 11 | 1.47 | 6.13 | 173 | 27 | 2.21 | 8.66 |
| Outpatient | 87 | 52 | 3.20 | 11.24 | 89 | 47 | 2.09 | 3.42 | 176 | 99 | 2.64 | 8.29 |
| Day patient | 81 | 6 | 0.10 | 0.48 | 87 | 5 | 0.07 | 0.37 | 168 | 11 | 0.08 | 0.43 |
| Accident and emergency | 87 | 20 | 0.22 | 0.42 | 91 | 11 | 0.12 | 0.32 | 178 | 31 | 0.17 | 0.38 |
| **NHS community-based services** |  |  |  |  |  |  |  |  |  |  |  |  |
| Intermediate care | 79 | 28 | 2.32 | 5.85 | 80 | 17 | 1.29 | 5.74 | 159 | 45 | 1.80 | 5.80 |
| General practitioner | 86 | 74 | 3.86 | 3.45 | 87 | 69 | 3.92 | 5.50 | 173 | 143 | 3.89 | 4.56 |
| General practitioner (Home) | 81 | 7 | 0.14 | 0.68 | 86 | 5 | 0.03 | 0.18 | 167 | 12 | 0.09 | 0.50 |
| Practice nurse | 80 | 44 | 1.17 | 1.93 | 85 | 36 | 0.78 | 1.32 | 165 | 80 | 0.97 | 1.65 |
| Practice nurse (Home) | 79 | 11 | 0.74 | 3.48 | 85 | 4 | 0.14 | 1.10 | 164 | 15 | 0.43 | 2.56 |
| Therapy assistant | 80 | 3 | 0.06 | 0.45 | 89 | 8 | 0.11 | 0.46 | 169 | 11 | 0.09 | 0.46 |
| Therapy assistant (Home) | 83 | 9 | 0.66 | 3.47 | 83 | 10 | 0.26 | 0.94 | 166 | 19 | 0.46 | 2.56 |
| Health care support worker | 77 | 4 | 0.10 | 0.52 | 85 | 2 | 0.03 | 0.24 | 162 | 6 | 0.07 | 0.40 |
| Health care support worker (Home) | 80 | 10 | 0.29 | 1.13 | 85 | 6 | 0.56 | 4.60 | 165 | 16 | 0.43 | 3.38 |
| Community nurse | 82 | 23 | 1.61 | 7.69 | 87 | 15 | 0.23 | 0.97 | 169 | 38 | 0.91 | 5.46 |
| Geriatrician or psychiatrist | 81 | 1 | 0.00 | 0.00 | 84 | 2 | 0.05 | 0.30 | 165 | 3 | 0.02 | 0.22 |
| Psychologist | 81 | 4 | 0.10 | 0.64 | 84 | 2 | 0.05 | 0.43 | 165 | 6 | 0.07 | 0.54 |
| Chiropodist | 81 | 12 | 0.61 | 2.52 | 85 | 11 | 0.23 | 0.89 | 166 | 23 | 0.42 | 1.90 |
| Chiropractor | 80 | 1 | 0.00 | 0.00 | 84 | 0 | 0.00 | 0.00 | 164 | 1 | 0.00 | 0.00 |
| Osteopath | 80 | 1 | 0.00 | 0.00 | 83 | 0 | 0.00 | 0.00 | 163 | 1 | 0.00 | 0.00 |
| Dentist | 83 | 22 | 0.35 | 0.78 | 84 | 25 | 0.35 | 0.83 | 167 | 47 | 0.35 | 0.80 |
| Optician | 81 | 35 | 0.53 | 0.79 | 85 | 31 | 0.58 | 1.15 | 166 | 66 | 0.56 | 0.98 |
| Complementary therapist | 80 | 3 | 0.05 | 0.44 | 83 | 2 | 0.00 | 0.00 | 163 | 5 | 0.02 | 0.31 |
| Day hospital | 81 | 2 | 0.01 | 0.11 | 85 | 4 | 0.00 | 0.00 | 166 | 6 | 0.01 | 0.08 |
| **Non-NHS community-based services** |  |  |  |  |  |  |  |  |  |  |  |  |
| Social club | 80 | 3 | 0.10 | 0.88 | 84 | 7 | 0.22 | 1.41 | 164 | 10 | 0.16 | 1.17 |
| Lunch club | 81 | 2 | 0.02 | 0.22 | 84 | 3 | 0.04 | 0.33 | 165 | 5 | 0.03 | 0.28 |
| Drop-in centre | 78 | 2 | 0.02 | 0.22 | 83 | 2 | 0.00 | 0.00 | 161 | 4 | 0.01 | 0.16 |
| Meals on wheels | 84 | 0 | 0.00 | 0.00 | 88 | 0 | 0.00 | 0.00 | 172 | 0 | 0.00 | 0.00 |
| Frozen meals on wheels | 84 | 1 | 0.00 | 0.00 | 88 | 0 | 0.00 | 0.00 | 172 | 1 | 0.00 | 0.00 |
| Home help for personal care | 81 | 9 | 0.72 | 4.96 | 89 | 5 | 0.83 | 5.39 | 170 | 14 | 0.78 | 5.18 |
| Home help for household tasks | 82 | 5 | 0.61 | 3.69 | 87 | 6 | 0.29 | 1.70 | 169 | 11 | 0.45 | 2.86 |
| Home help for shopping assistance | 80 | 1 | 0.00 | 0.00 | 88 | 2 | 0.00 | 0.00 | 168 | 3 | 0.00 | 0.00 |
| Social worker | 83 | 8 | 0.16 | 0.67 | 87 | 1 | 0.01 | 0.11 | 170 | 9 | 0.09 | 0.48 |
| Social worker (Telephone) | 82 | 4 | 0.08 | 0.44 | 86 | 2 | 0.02 | 0.21 | 168 | 6 | 0.05 | 0.35 |
| Day care centre | 81 | 0 | 0.00 | 0.00 | 86 | 0 | 0.00 | 0.00 | 167 | 0 | 0.00 | 0.00 |
| Residential care home | 83 | 3 | 1.43 | 8.15 | 90 | 0 | 0.00 | 0.00 | 173 | 3 | 0.69 | 5.71 |
| **Therapy services in the community** |  |  |  |  |  |  |  |  |  |  |  |  |
| Speech and language therapy | 71 | 6 | 0.09 | 0.41 | 81 | 8 | 0.13 | 0.76 | 152 | 14 | 0.11 | 0.61 |
| Speech and language therapy (Home) | 78 | 18 | 0.78 | 3.24 | 80 | 11 | 0.20 | 0.96 | 158 | 29 | 0.49 | 2.41 |
| Physiotherapy | 68 | 10 | 0.46 | 1.53 | 84 | 7 | 0.24 | 1.07 | 152 | 17 | 0.34 | 1.30 |
| Physiotherapy (Home) | 79 | 50 | 5.62 | 11.33 | 88 | 34 | 1.98 | 4.52 | 167 | 84 | 3.64 | 8.52 |
| Occupational therapy | 69 | 15 | 0.34 | 0.92 | 82 | 16 | 0.29 | 0.87 | 151 | 31 | 0.31 | 0.89 |
| Occupational therapy (Home) | 79 | 38 | 2.08 | 4.33 | 88 | 35 | 1.99 | 9.23 | 167 | 73 | 2.03 | 7.22 |
| **Unpaid help** |  |  |  |  |  |  |  |  |  |  |  |  |
| Help from people living with participants | 84 | 45 | 146.54 | 431.52 | 87 | 33 | 64.29 | 183.37 | 171 | 78 | 103.70 | 328.10 |
| Help from people not living with participants | 79 | 23 | 28.36 | 104.13 | 84 | 25 | 15.45 | 95.46 | 163 | 48 | 21.69 | 99.62 |

**Table S3:** Resource use: number of participants using a service and mean (SD) contacts at 12-month (for the previous 6 months)

|  | GSG (N=180) |  |  |  | Usual care (N=151) |  |  |  | Overall (N=331) |  |  |  |
| --- | --- | --- | --- | --- | --- | --- | --- | --- | --- | --- | --- | --- |
| Resource | (N) returned | (N) used | (Mean) | (SD) | (N) returned | (N) used | (Mean) | (SD) | (N) returned | (N) used | (Mean) | (SD) |
| **Hospital services** |  |  |  |  |  |  |  |  |  |  |  |  |
| Hospital admission | 88 | 12 | 0.61 | 3.05 | 90 | 14 | 0.74 | 2.61 | 178 | 26 | 0.67 | 2.83 |
| Outpatient | 89 | 39 | 1.44 | 3.64 | 90 | 43 | 1.46 | 2.53 | 179 | 82 | 1.45 | 3.14 |
| Day patient | 88 | 6 | 0.08 | 0.35 | 88 | 9 | 0.11 | 0.44 | 176 | 15 | 0.10 | 0.39 |
| Accident and emergency | 88 | 13 | 0.14 | 0.34 | 91 | 17 | 0.18 | 0.39 | 179 | 30 | 0.16 | 0.37 |
| **NHS Community-based services** |  |  |  |  |  |  |  |  |  |  |  |  |
| Intermediate care | 76 | 8 | 0.94 | 5.41 | 83 | 7 | 0.08 | 0.39 | 159 | 15 | 0.51 | 3.85 |
| General practitioner | 83 | 68 | 4.23 | 5.24 | 87 | 70 | 3.45 | 3.75 | 170 | 138 | 3.86 | 4.60 |
| General practitioner (Home) | 78 | 8 | 0.08 | 0.32 | 84 | 3 | 0.06 | 0.35 | 162 | 11 | 0.07 | 0.34 |
| Practice nurse | 80 | 38 | 0.96 | 1.51 | 86 | 33 | 0.59 | 1.19 | 166 | 71 | 0.78 | 1.37 |
| Practice nurse (Home) | 79 | 3 | 0.06 | 0.39 | 85 | 5 | 0.03 | 0.18 | 164 | 8 | 0.05 | 0.30 |
| Therapy assistant | 79 | 1 | 0.01 | 0.11 | 85 | 5 | 0.06 | 0.35 | 164 | 6 | 0.03 | 0.26 |
| Therapy assistant (Home) | 79 | 4 | 0.15 | 1.02 | 85 | 3 | 0.03 | 0.32 | 164 | 7 | 0.09 | 0.75 |
| Health care support worker | 78 | 0 | 0.00 | 0.00 | 86 | 3 | 0.05 | 0.30 | 164 | 3 | 0.02 | 0.21 |
| Health care support worker (Home) | 79 | 6 | 1.43 | 10.51 | 84 | 4 | 0.13 | 0.78 | 163 | 10 | 0.78 | 7.48 |
| Community nurse | 79 | 11 | 0.32 | 1.30 | 83 | 4 | 0.01 | 0.11 | 162 | 15 | 0.17 | 0.94 |
| Geriatrician or psychiatrist | 81 | 0 | 0.00 | 0.00 | 86 | 3 | 0.00 | 0.00 | 167 | 3 | 0.00 | 0.00 |
| Psychologist | 80 | 2 | 0.02 | 0.21 | 86 | 6 | 0.12 | 1.09 | 166 | 8 | 0.07 | 0.78 |
| Chiropodist | 80 | 14 | 0.34 | 0.99 | 84 | 13 | 0.31 | 0.97 | 164 | 27 | 0.33 | 0.98 |
| Chiropractor | 81 | 2 | 0.03 | 0.32 | 82 | 2 | 0.00 | 0.00 | 163 | 4 | 0.02 | 0.23 |
| Osteopath | 81 | 4 | 0.07 | 0.48 | 84 | 2 | 0.00 | 0.00 | 165 | 6 | 0.03 | 0.34 |
| Dentist | 82 | 30 | 0.49 | 0.92 | 86 | 36 | 0.54 | 0.88 | 168 | 66 | 0.51 | 0.90 |
| Optician | 81 | 27 | 0.39 | 0.75 | 85 | 31 | 0.52 | 0.99 | 166 | 58 | 0.45 | 0.87 |
| Complementary therapist | 80 | 3 | 0.15 | 1.29 | 84 | 2 | 0.00 | 0.00 | 164 | 5 | 0.08 | 0.92 |
| Day hospital | 79 | 3 | 0.06 | 0.39 | 84 | 4 | 0.05 | 0.44 | 163 | 7 | 0.05 | 0.41 |
| **Non-NHS Community-based services** |  |  |  |  |  |  |  |  |  |  |  |  |
| Social club | 78 | 4 | 0.04 | 0.24 | 85 | 4 | 0.02 | 0.22 | 163 | 8 | 0.03 | 0.23 |
| Lunch club | 79 | 4 | 0.06 | 0.45 | 83 | 4 | 0.02 | 0.22 | 162 | 8 | 0.04 | 0.35 |
| Drop-in centre | 77 | 1 | 0.00 | 0.00 | 84 | 3 | 0.02 | 0.22 | 161 | 4 | 0.01 | 0.15 |
| Meals on wheels | 84 | 1 | 0.00 | 0.00 | 88 | 0 | 0.00 | 0.00 | 172 | 1 | 0.00 | 0.00 |
| Frozen meals on wheels | 84 | 2 | 0.13 | 1.26 | 88 | 1 | 0.26 | 2.52 | 172 | 3 | 0.20 | 1.98 |
| Home help for personal care | 85 | 5 | 1.46 | 10.73 | 88 | 3 | 2.24 | 18.96 | 173 | 8 | 1.85 | 15.39 |
| Home help for household tasks | 84 | 2 | 0.09 | 0.84 | 87 | 6 | 0.61 | 3.38 | 171 | 8 | 0.35 | 2.45 |
| Home help for shopping assistance | 84 | 2 | 0.00 | 0.00 | 87 | 0 | 0.00 | 0.00 | 171 | 2 | 0.00 | 0.00 |
| Social worker | 82 | 2 | 0.01 | 0.11 | 87 | 1 | 0.00 | 0.00 | 169 | 3 | 0.01 | 0.07 |
| Social worker (Telephone) | 83 | 1 | 0.00 | 0.00 | 87 | 0 | 0.00 | 0.00 | 170 | 1 | 0.00 | 0.00 |
| Day care centre | 82 | 1 | 0.00 | 0.00 | 88 | 0 | 0.00 | 0.00 | 170 | 1 | 0.00 | 0.00 |
| Residential care home | 87 | 4 | 6.33 | 41.44 | 89 | 2 | 0.08 | 0.73 | 176 | 6 | 3.27 | 29.71 |
| **Therapy services in the community** |  |  |  |  |  |  |  |  |  |  |  |  |
| Speech and language therapy | 83 | 2 | 0.07 | 0.63 | 87 | 3 | 0.11 | 1.07 | 170 | 5 | 0.09 | 0.87 |
| Speech and language therapy (Home) | 84 | 4 | 0.30 | 2.55 | 84 | 2 | 0.00 | 0.00 | 168 | 6 | 0.15 | 1.83 |
| Physiotherapy | 81 | 10 | 0.64 | 4.33 | 87 | 10 | 0.61 | 2.99 | 168 | 20 | 0.63 | 3.71 |
| Physiotherapy (Home) | 84 | 13 | 0.38 | 1.38 | 86 | 11 | 0.47 | 2.76 | 170 | 24 | 0.42 | 2.16 |
| Occupational therapy | 82 | 7 | 0.69 | 5.15 | 85 | 4 | 0.01 | 0.11 | 167 | 11 | 0.36 | 3.68 |
| Occupational therapy (Home) | 83 | 9 | 0.09 | 0.39 | 85 | 7 | 0.06 | 0.36 | 168 | 16 | 0.08 | 0.37 |
| **Unpaid help** |  |  |  |  |  |  |  |  |  |  |  |  |
| Help from people living with participants | 86 | 36 | 121.57 | 537.67 | 86 | 26 | 77.76 | 392.81 | 172 | 62 | 99.06 | 467.70 |
| Help from people not living with participants | 81 | 15 | 11.36 | 67.24 | 85 | 17 | 10.43 | 38.51 | 166 | 32 | 10.89 | 54.36 |

**Table S4:** Resource use number of participants using a service and mean (SD) contacts at 24-month (for the previous 6 months)

|  | **GSG**<br>**(N=180)** |  |  |  | **Usual care**<br>**(N=151)** |  |  |  | **Overall**<br>**(N=331)** |  |  |  |
| --- | --- | --- | --- | --- | --- | --- | --- | --- | --- | --- | --- | --- |
| **Resource** | **(N)**<br>**returned** | **(N) used** | **(Mean)** | **(SD)** | **(N)**<br>**returned** | **(N) used** | **(Mean)** | **(SD)** | **(N) returned** | **(N) used** | **(Mean)** | **(SD)** |
| ***Hospital services*** |  |  |  |  |  |  |  |  |  |  |  |  |
| Hospital admission | 28 | 2 | 0.26 | 1.18 | 29 | 3 | 0.16 | 0.57 | 57 | 5 | 0.21 | 0.96 |
| Outpatient | 26 | 14 | 1.05 | 2.54 | 29 | 15 | 1.32 | 1.70 | 55 | 29 | 1.18 | 2.19 |
| Day patient | 27 | 5 | 0.14 | 0.42 | 29 | 2 | 0.21 | 0.73 | 56 | 7 | 0.17 | 0.57 |
| Accident and emergency | 28 | 3 | 0.07 | 0.26 | 29 | 4 | 0.13 | 0.34 | 57 | 7 | 0.09 | 0.29 |
| ***NHS Community based services*** |  |  |  |  |  |  |  |  |  |  |  |  |
| Intermediate care | 24 | 2 | 0.54 | 3.20 | 26 | 1 | 0.14 | 0.74 | 50 | 3 | 0.37 | 2.47 |
| General practitioner | 28 | 19 | 1.79 | 2.67 | 28 | 22 | 2.57 | 2.85 | 56 | 41 | 2.10 | 2.75 |
| General practitioner (Home) | 28 | 1 | 0.02 | 0.15 | 28 | 2 | 0.03 | 0.18 | 56 | 3 | 0.03 | 0.16 |
| Practice nurse | 27 | 6 | 0.26 | 0.70 | 27 | 12 | 0.57 | 0.84 | 54 | 18 | 0.39 | 0.77 |
| Practice nurse (Home) | 28 | 0 | 0.00 | 0.00 | 28 | 1 | 0.00 | 0.00 | 56 | 1 | 0.00 | 0.00 |
| Therapy assistant | 27 | 1 | 0.02 | 0.15 | 27 | 1 | 0.00 | 0.00 | 54 | 2 | 0.01 | 0.12 |
| Therapy assistant (Home) | 28 | 0 | 0.00 | 0.00 | 27 | 1 | 0.00 | 0.00 | 55 | 1 | 0.00 | 0.00 |
| Health care support worker | 28 | 1 | 0.00 | 0.00 | 28 | 2 | 0.07 | 0.37 | 56 | 3 | 0.03 | 0.24 |
| Health care support worker (Home) | 28 | 1 | 0.47 | 3.05 | 28 | 1 | 0.00 | 0.00 | 56 | 2 | 0.27 | 2.34 |
| Community nurse | 26 | 0 | 0.00 | 0.00 | 27 | 1 | 0.00 | 0.00 | 53 | 1 | 0.00 | 0.00 |
| Geriatrician or psychiatrist | 26 | 1 | 0.02 | 0.16 | 27 | 0 | 0.00 | 0.00 | 53 | 1 | 0.01 | 0.12 |
| Psychologist | 26 | 0 | 0.00 | 0.00 | 28 | 1 | 0.00 | 0.00 | 54 | 1 | 0.00 | 0.00 |
| Chiropodist | 27 | 5 | 0.31 | 0.98 | 27 | 3 | 0.27 | 0.83 | 54 | 8 | 0.29 | 0.91 |
| Chiropractor | 26 | 1 | 0.02 | 0.16 | 27 | 1 | 0.00 | 0.00 | 53 | 2 | 0.01 | 0.12 |
| Osteopath | 26 | 0 | 0.00 | 0.00 | 25 | 0 | 0.00 | 0.00 | 51 | 0 | 0.00 | 0.00 |
| Dentist | 26 | 9 | 0.27 | 0.55 | 27 | 16 | 1.00 | 1.73 | 53 | 25 | 0.56 | 1.21 |
| Optician | 26 | 5 | 0.15 | 0.42 | 29 | 12 | 0.45 | 0.78 | 55 | 17 | 0.27 | 0.61 |
| Complementary therapist | 26 | 0 | 0.00 | 0.00 | 26 | 2 | 0.07 | 0.38 | 52 | 2 | 0.03 | 0.24 |
| Day hospital | 27 | 0 | 0.00 | 0.00 | 28 | 2 | 0.13 | 0.73 | 55 | 2 | 0.06 | 0.47 |
| **Non-NHS community-based services** |  |  |  |  |  |  |  |  |  |  |  |  |
| Social club | 26 | 1 | 0.00 | 0.00 | 27 | 3 | 0.00 | 0.00 | 53 | 4 | 0.00 | 0.00 |
| Lunch club | 26 | 1 | 0.15 | 0.94 | 26 | 1 | 0.00 | 0.00 | 52 | 2 | 0.09 | 0.72 |
| Drop-in centre | 25 | 0 | 0.00 | 0.00 | 27 | 1 | 0.00 | 0.00 | 52 | 1 | 0.00 | 0.00 |
| Meals on wheels | 28 | 0 | 0.00 | 0.00 | 28 | 0 | 0.00 | 0.00 | 56 | 0 | 0.00 | 0.00 |
| Frozen meals on wheels | 28 | 1 | 0.09 | 0.61 | 28 | 0 | 0.00 | 0.00 | 56 | 1 | 0.05 | 0.46 |
| Home help for personal care | 28 | 0 | 0.00 | 0.00 | 28 | 0 | 0.00 | 0.00 | 56 | 0 | 0.00 | 0.00 |
| Home help for household tasks | 28 | 1 | 0.00 | 0.00 | 27 | 0 | 0.00 | 0.00 | 55 | 1 | 0.00 | 0.00 |
| Home help for shopping assistance | 28 | 0 | 0.00 | 0.00 | 28 | 0 | 0.00 | 0.00 | 56 | 0 | 0.00 | 0.00 |
| Social worker | 28 | 0 | 0.00 | 0.00 | 29 | 1 | 0.00 | 0.00 | 57 | 1 | 0.00 | 0.00 |
| Social worker (Telephone) | 28 | 0 | 0.00 | 0.00 | 29 | 2 | 0.03 | 0.18 | 57 | 2 | 0.01 | 0.12 |
| Day care centre | 28 | 0 | 0.00 | 0.00 | 28 | 0 | 0.00 | 0.00 | 56 | 0 | 0.00 | 0.00 |
| Residential care home | 27 | 0 | 0.00 | 0.00 | 28 | 0 | 0.00 | 0.00 | 55 | 0 | 0.00 | 0.00 |
| **Therapy services in the community** |  |  |  |  |  |  |  |  |  |  |  |  |
| Speech and Language Therapy | 28 | 1 | 0.00 | 0.00 | 32 | 0 | 0.00 | 0.00 | 60 | 1 | 0.00 | 0.00 |
| Speech and Language Therapy (Home) | 27 | 0 | 0.00 | 0.00 | 30 | 0 | 0.00 | 0.00 | 57 | 0 | 0.00 | 0.00 |
| Physiotherapy | 28 | 2 | 0.05 | 0.21 | 33 | 4 | 0.34 | 1.35 | 61 | 6 | 0.18 | 0.92 |
| Physiotherapy (Home) | 28 | 0 | 0.00 | 0.00 | 32 | 0 | 0.00 | 0.00 | 60 | 0 | 0.00 | 0.00 |
| Occupational Therapy | 27 | 1 | 0.00 | 0.00 | 33 | 2 | 0.64 | 3.36 | 60 | 3 | 0.30 | 2.30 |
| Occupational Therapy (Home) | 28 | 0 | 0.00 | 0.00 | 32 | 0 | 0.00 | 0.00 | 60 | 0 | 0.00 | 0.00 |
| **Unpaid Help** |  |  |  |  |  |  |  |  |  |  |  |  |
| Help from people living with participant | 28 | 9 | 32.93 | 139.86 | 34 | 13 | 185.69 | 429.14 | 62 | 22 | 99.89 | 309.81 |
| Help from people not living with participant | 23 | 1 | 0.84 | 5.19 | 32 | 6 | 6.41 | 24.09 | 55 | 7 | 3.39 | 16.82 |

**Table S5:** Costs: number of participants using a service and mean (SD) cost per participant at baseline (for the previous 3 months)

|  | GSG (N=180) |  |  | Usual care (N=151) |  |  | Overall (N=331) |  |  |
| --- | --- | --- | --- | --- | --- | --- | --- | --- | --- |
| Resources | (N) | (Mean) | (SD) | (N) | (Mean) | (SD) | (N) | (Mean) | (SD) |
| Hospital admissions | 168 | 289.8 | 1341.2 | 150 | 255.1 | 1146.6 | 318 | 273.4 | 1251.3 |
| Outpatient visits | 167 | 98.7 | 209.6 | 148 | 93.1 | 318.2 | 315 | 96.1 | 265.8 |
| Day patient visits | 165 | 54.2 | 255.5 | 145 | 6.2 | 74.2 | 310 | 31.7 | 194.4 |
| Accident and emergency visits | 169 | 48.9 | 165.9 | 147 | 35.8 | 134.0 | 316 | 42.8 | 151.8 |
| **Total Hospital Services** | 160 | 457.6 | 1493.8 | 140 | 377.4 | 1303.3 | 300 | 420.2 | 1406.4 |
| Therapy services in the community | 171 | 10.9 | 57.3 | 151 | 20.4 | 115.8 | 322 | 15.4 | 89.6 |
| NHS community-based services | 120 | 148.7 | 171.6 | 90 | 137.1 | 202.1 | 210 | 143.7 | 184.9 |
| Non-NHS community-based services | 122 | 22.8 | 84.2 | 125 | 30.2 | 154.9 | 247 | 26.5 | 124.9 |
| Unpaid help | 172 | 2672.1 | 6248.5 | 150 | 1532.1 | 4853.5 | 322 | 2141.1 | 5662.0 |
| **Total NHS/PSS<sup>1</sup>** | 115 | 657.6 | 1530.9 | 85 | 487.3 | 1172.8 | 200 | 585.2 | 1389.4 |
| **Total societal<sup>2</sup>** | 89 | 2710.2 | 4222.1 | 70 | 1452.1 | 2812.4 | 159 | 2156.3 | 3711.5 |

<sup>1</sup>Total NHS/PSS cost consists of total hospital services cost plus therapy services in the community and NHS community-based services costs
<sup>2</sup>Total Societal cost consists of total NHS cost plus non-NHS community-based services and unpaid help costs

**Table S6:** Costs: number of participants using a service and mean (SD) cost per participant at 6-months (for the previous 6 months)

|  | GSG (N=180) |  |  | Usual care (N=151) |  |  | Overall (N=331) |  |  |
| --- | --- | --- | --- | --- | --- | --- | --- | --- | --- |
| Resources | (N) | (Mean) | (SD) | (N) | (Mean) | (SD) | (N) | (Mean) | (SD) |
| Hospital admissions | 85 | 736.5 | 1705.1 | 93 | 635.8 | 2232.1 | 178 | 683.9 | 1993.0 |
| Outpatient visits | 91 | 318.6 | 566.3 | 91 | 399.2 | 789.6 | 182 | 358.9 | 686.4 |
| Day patient visits | 84 | 85.1 | 430.4 | 87 | 61.7 | 328.1 | 171 | 73.2 | 380.9 |
| Accident and emergency visits | 91 | 115.3 | 259.4 | 93 | 59.1 | 182.2 | 184 | 86.9 | 224.9 |
| **Total hospital services** | 81 | 1124.8 | 2103.6 | 84 | 866.9 | 1620.4 | 165 | 993.5 | 1871.9 |
| Therapy services in the community | 72 | 841.9 | 1687.6 | 86 | 390.7 | 962.9 | 158 | 596.3 | 1356.8 |
| NHS community-based services | 51 | 492.2 | 698.5 | 49 | 379.3 | 411.7 | 100 | 436.9 | 576.0 |
| Non-NHS community-based services | 55 | 197.3 | 1179.8 | 66 | 29.8 | 118.9 | 121 | 105.9 | 800.6 |
| Unpaid help | 90 | 16715.4 | 29434.7 | 94 | 10309.8 | 23248.4 | 184 | 13443.0 | 26577.2 |
| **Total NHS/PSS<sup>1</sup>** | 39 | 2086.7 | 2626.9 | 45 | 1689.6 | 1974.8 | 84 | 1873.9 | 2294.9 |
| **Total societal<sup>2</sup>** | 31 | 20120.5 | 31592.6 | 39 | 12081.9 | 24069.8 | 70 | 15641.8 | 27734.3 |

<sup>1</sup>Total NHS /PSS cost consists of total hospital services cost plus therapy services in the community and NHS community-based services costs
<sup>2</sup>Total Societal cost consists of total NHS cost plus non-NHS community-based services and unpaid help costs

**Table S7:**
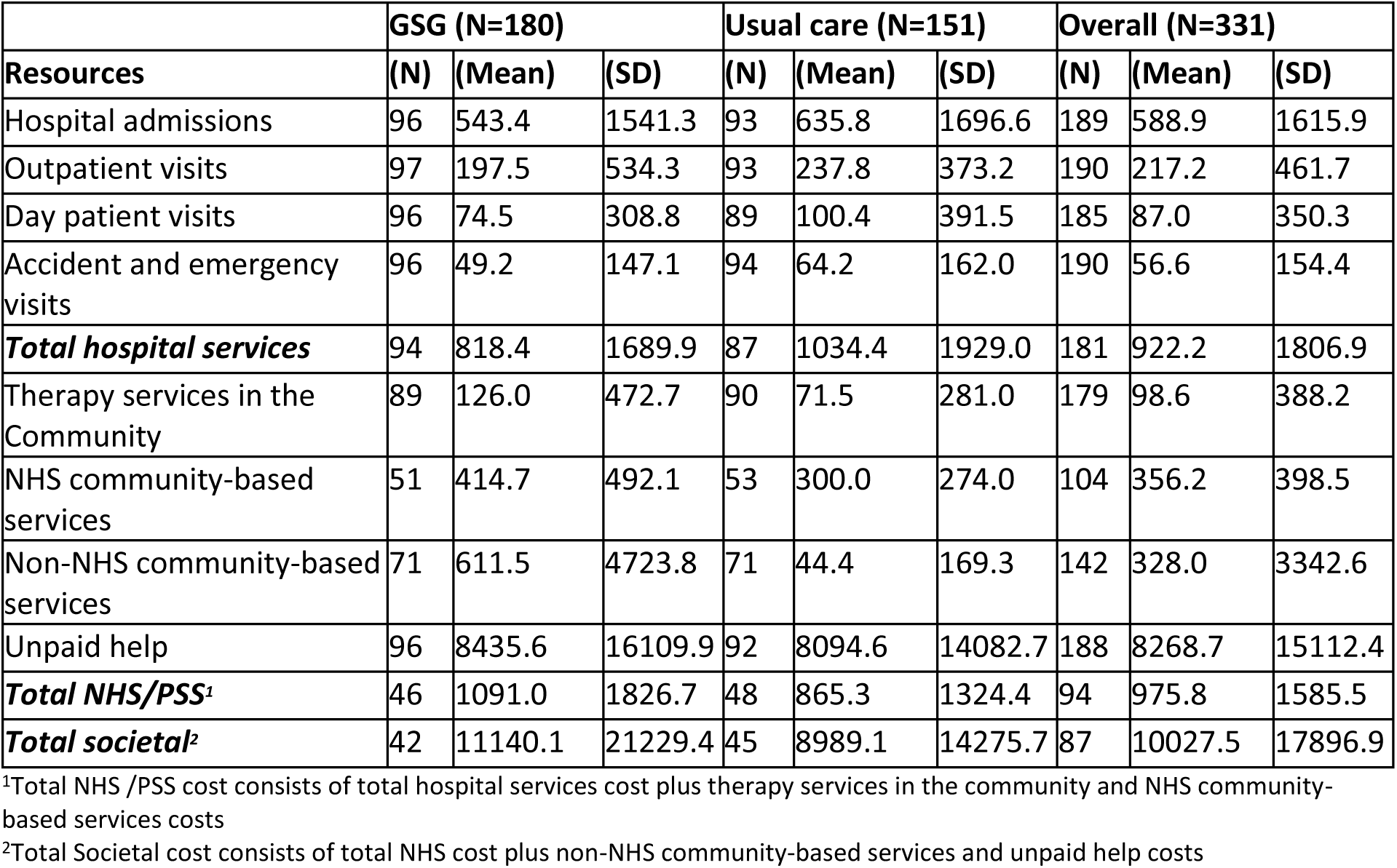
Costs: number of participants using a service and mean (SD) cost per participant at 12-months (for the previous 6 months)

**Table S8:**
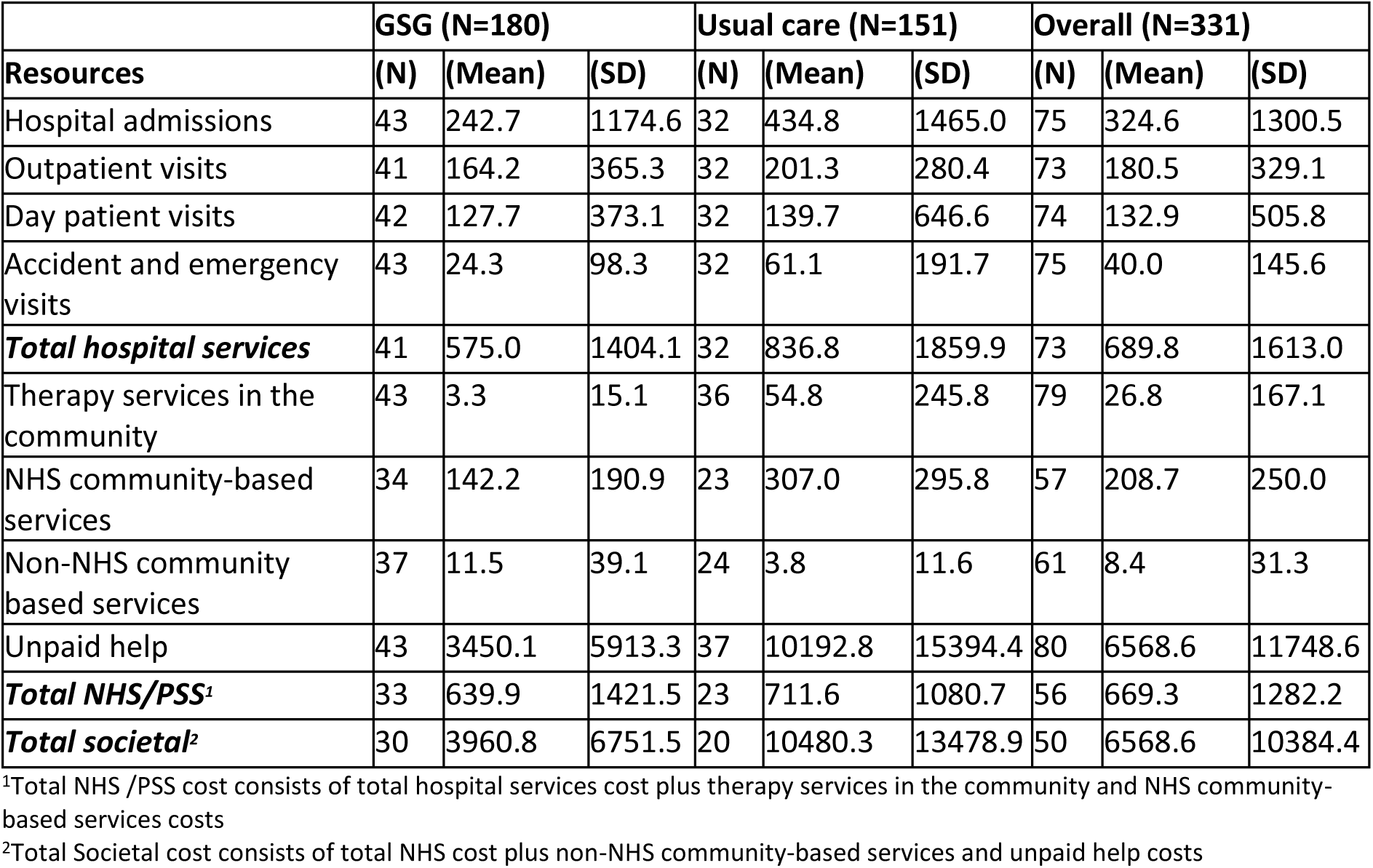
Costs: number of participants using a service and mean (SD) cost per participant at 24-months (for the previous 6 months)

|  | GSG (N=180) |  |  | Usual care (N=151) |  |  | Overall (N=331) |  |  |
| --- | --- | --- | --- | --- | --- | --- | --- | --- | --- |
| **Resources** | **(N)** | **(Mean)** | **(SD)** | **(N)** | **(Mean)** | **(SD)** | **(N)** | **(Mean)** | **(SD)** |
| Hospital admissions | 43 | 242.7 | 1174.6 | 32 | 434.8 | 1465.0 | 75 | 324.6 | 1300.5 |
| Outpatient visits | 41 | 164.2 | 365.3 | 32 | 201.3 | 280.4 | 73 | 180.5 | 329.1 |
| Day patient visits | 42 | 127.7 | 373.1 | 32 | 139.7 | 646.6 | 74 | 132.9 | 505.8 |
| Accident and emergency visits | 43 | 24.3 | 98.3 | 32 | 61.1 | 191.7 | 75 | 40.0 | 145.6 |
| **Total hospital services** | 41 | 575.0 | 1404.1 | 32 | 836.8 | 1859.9 | 73 | 689.8 | 1613.0 |
| Therapy services in the community | 43 | 3.3 | 15.1 | 36 | 54.8 | 245.8 | 79 | 26.8 | 167.1 |
| NHS community-based services | 34 | 142.2 | 190.9 | 23 | 307.0 | 295.8 | 57 | 208.7 | 250.0 |
| Non-NHS community based services | 37 | 11.5 | 39.1 | 24 | 3.8 | 11.6 | 61 | 8.4 | 31.3 |
| Unpaid help | 43 | 3450.1 | 5913.3 | 37 | 10192.8 | 15394.4 | 80 | 6568.6 | 11748.6 |
| **Total NHS/PSS<sup>1</sup>** | 33 | 639.9 | 1421.5 | 23 | 711.6 | 1080.7 | 56 | 669.3 | 1282.2 |
| **Total societal<sup>2</sup>** | 30 | 3960.8 | 6751.5 | 20 | 10480.3 | 13478.9 | 50 | 6568.6 | 10384.4 |

<sup>1</sup>Total NHS /PSS cost consists of total hospital services cost plus therapy services in the community and NHS community-based services costs
<sup>2</sup>Total Societal cost consists of total NHS cost plus non-NHS community-based services and unpaid help costs

**Figure S1.**
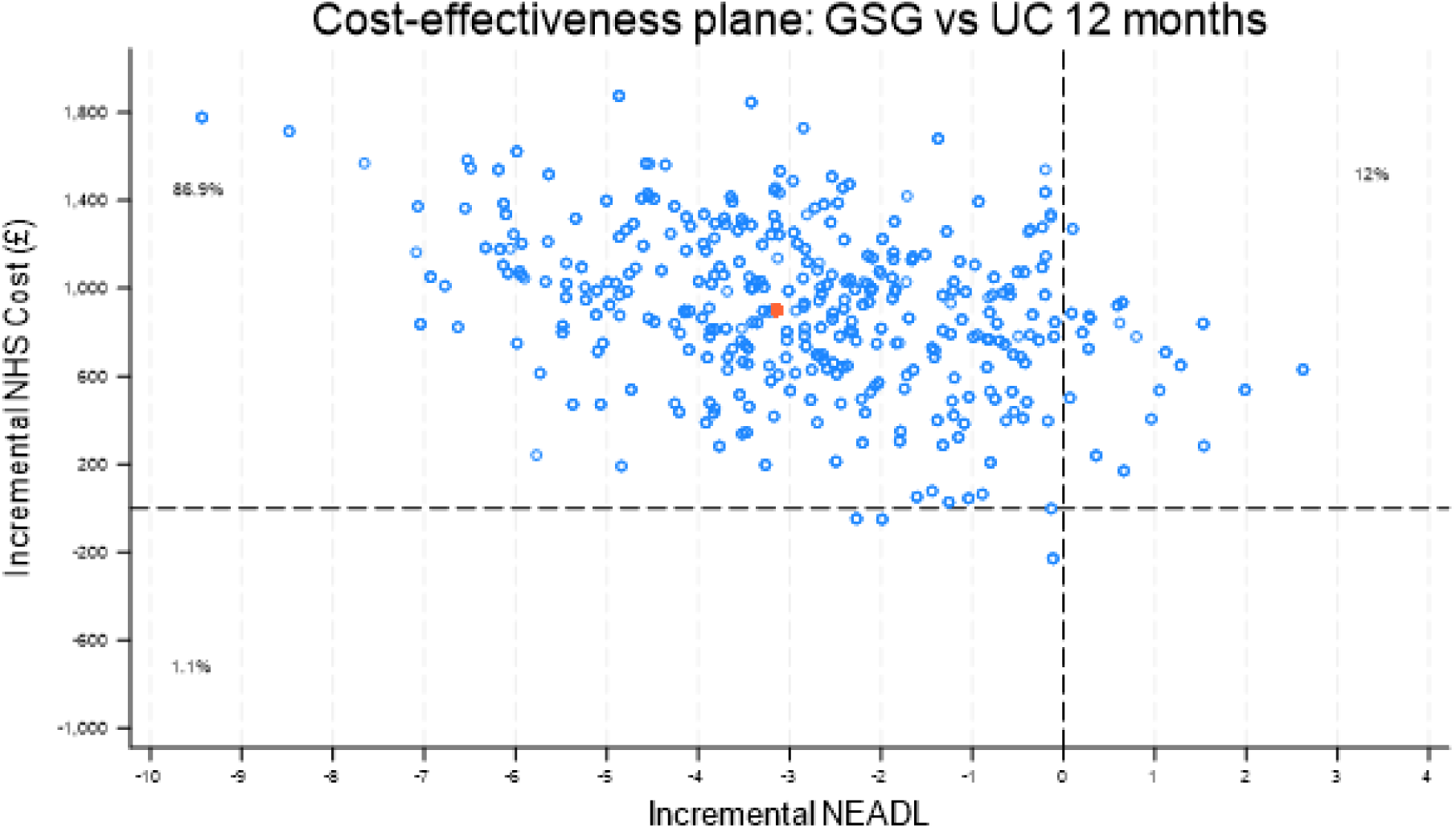
Cost-effectiveness plane for costs from NHS & PSS perspective and NEADL differences.

